# Peru’s front-of-package warning label policy and changes in the healthfulness of the food supply: A pre-post observational study

**DOI:** 10.1101/2025.05.20.25328034

**Authors:** Jonathan Lara-Arevalo, Lorena Saavedra-Garcia, Francisco Diez-Canseco, Natalia Rebolledo, Donna R. Miles, Bridget Hollingsworth, J. Jaime Miranda, Lindsey Smith Taillie

**Affiliations:** Department of Nutrition, Gillings School of Global Public Health, University of North Carolina, Chapel Hill, North Carolina, USA; CRONICAS Centre of Excellence in Chronic Diseases, Universidad Peruana Cayetano Heredia, Lima, Peru; Institute of Nutrition and Food Technology (INTA), University of Chile, Macul, Metropolitan Region, Chile; Carolina Population Center, University of North Carolina, Chapel Hill, North Carolina, USA

## Abstract

**Background:** In June 2019, Peru implemented the first phase of a front-of-package warning label (FOPWL) policy, requiring packaged foods and beverages exceeding established thresholds for total sugars, sodium, saturated fats, or containing trans fats to display warning labels. While primarily intended to inform consumers, such policies may also incentivize product reformulation. This study aimed to assess changes in the nutrient content and proportion of regulated products (i.e., “high in” products) in the Peruvian food supply following the implementation of the first phase of the policy.

**Methods and Findings:** A dataset of nutritional information from 2019 to 2021 was developed using product-level data collected at two time periods: preimplementation (T0: March–June 2019; n = 2,481) and postimplementation (T1: March 2020–February 2021; n = 3,018). The proportion of products exceeding regulatory thresholds (per 100 g or 100 mL) for total sugars, saturated fats, sodium, or containing trans fats, as well as quartiles of these nutrients, were compared pre-and postimplementation of the law in cross-sectional samples of products. A longitudinal subsample (i.e., products collected in both the pre-and postimplementation periods, n = 1,694) was also analyzed. Changes in the proportion of “high in” products were assessed using logistic regression models; shifts in nutrient distributions were evaluated using quantile regression models. In cross-sectional analyses over T1-T0 period, the share of “high in” products declined by 6 percentage points, from 62% to 56% among foods and by 11 percentage points, from 32% to 21% among beverages. Relative reductions were observed in products “high in” sugars (10% for foods; 36% for beverages), sodium (19% for foods), and trans fats (41% for foods). The reductions varied across food and beverage categories and were greatest in groups where the regulatory thresholds were set below the 75th percentile of the baseline nutrient distributions. Longitudinal analyses showed results largely consistent with the cross-sectional findings.

**Conclusions:** The first phase of Peru’s FOPWL policy was associated with substantial reductions in the proportion of products “high in” nutrients of concern, suggesting that the policy led to reformulation across diverse food and beverage categories. These findings highlight the potential of mandatory FOPWL policies to improve the nutritional quality of the food supply and support population-wide strategies for preventing diet-related diseases as modest percentage-point reductions in “high in” products can yield meaningful health improvements when applied across millions of consumers.

## Introduction

In recent decades, the global prevalence of obesity and diet-related noncommunicable diseases (NCDs) has risen sharply, particularly in low-and middle-income countries (LMICs), contributing substantially to the burden of related healthcare costs (1). In Peru, the prevalence of overweight and obesity in individuals over 15 years old increased from 53.2% to 61.3% between 2015 and 2023, with similar upward trends among children (2, 3). This increase in NCD risk is closely linked to dietary changes, particularly the rising consumption of sugar-sweetened beverages (SSBs) and ultra-processed foods (UPFs), which are often high in sugars, saturated fats, sodium, and other nutrients of concern (4–6). As UPFs and SSBs become more available and affordable in LMICs, their intake contributes to excess calorie consumption and increased cardiometabolic risk, further exacerbating the obesity and NCD crisis (7–9).

In response, many Latin American countries have implemented food policies to reduce UPFs and SSBs consumption (6). Peru’s Law No. 30021, “Promotion of Healthy Eating for Boys, Girls and Adolescents,” approved in 2013, is one such policy (10). This law, implemented in two phases (2019 and 2021, with stricter nutritional parameters in the second phase), mandates front-of-package warning labels (FOPWL) on UPFs and SSBs exceeding established thresholds for total sugar, saturated fats, sodium, or containing trans fats (10, 11). These labels, displayed as black octagons with white capital letters, alert consumers when a product is “high in” total sugar, saturated fats, or sodium, or if it “contains trans fats”. Additionally, they include supplementary text advising consumers to “avoid excessive consumption” or, in the case of trans fats, to “avoid its consumption” (10).

Although the primary goal of FOPWL policies like Peru’s typically is to inform consumers, growing evidence indicates that mandatory policies also incentivize the food industry to reduce or remove nutrients of concern through reformulation, product removal, or introduction of new products (12–14). Studies from Chile and Mexico, for instance, have documented industry reformulation efforts to reduce the content of nutrients of concern in UPFs and SSBs to avoid labeling (15–17). Understanding how similar policies influence the Peruvian food supply is especially relevant given the country’s unique regulatory landscape. Prior to implementing its FOPWL policy in 2019 and unlike other countries, Peru adopted two other measures targeting nutrients of concern: a 2016 mandate to progressively eliminate trans fats (18) and a 2018 increase in taxes on high-sugar beverages from 17% to 25%, aligned with FOPWL sugar thresholds (19). Peru’s FOPWL system also mandates warning labels across all advertising platforms—including television, social media, and radio—and was further strengthened in 2019 by Ministry of Health guidelines mandating the exclusion of products with FOPWL from school kiosks and cafeterias to reinforce healthy eating habits among children (10, 20). The combination of these consecutive regulatory actions positions Peru as a distinct case for evaluating industry responses to public health policies in a middle-income setting undergoing rapid dietary transitions.

This study aims to assess changes in the levels of sugar, sodium, saturated fats, and trans fats, as well as the prevalence of “high in” products and those contanining trans fats before and after the implementation of the first phase of Peru’s FOPWL law in 2019-2021.

## Methods

### Peruvian context: Peru’s FOPWL nutrient thresholds

Although initially approved in 2013, the FOPWL law was first implemented until 2019, due to significant opposition (21). It was implemented in two stages: the first phase, beginning in June 2019, introduced initial nutrient thresholds, while the second phase, in September 2021, imposed stricter nutritional limits to the same products (11). The cutoff values vary depending on whether the product is solid or liquid, and apply to the ready-to-consume version for products requiring reconstitution. For the first phase, cutoff values were as follows: total sugars—22.5 g per 100 g for solid foods and 6 g per 100 mL for liquids; sodium—800 mg per 100 g for solids and 100 mg per 100 mL for liquids; saturated fats—6 g per 100 g for solids and 3 g per 100 mL for liquids; and trans fats—any amount required labeling. The updated cutoffs for the second implementation phase are detailed in **S1 Table**.

### The Peruvian Nutrition Facts Panel (NFP) Database

A Nutrition Facts Panel (NFP) dataset was developed using data from 2019 to 2021 gathered from photographs of packaged food and beverages products in supermarkets in Lima, Peru, the capital and primary commercial center of the country. Data were obtained from three different establishments of a large national supermarket chain. Aiming to capture a large variety of packaged products, each selected supermarkets targeted one of three different socioeconomic sectors (high, medium, and low) thereby opening the possibility to find most of the products available in the Peruvian market. Data collection occurred at two distinct time points: pre-implementation (T0: 2019) and post-implementation (T1: 2020-2021) of the relevant law. In the first wave, data were collected during the pre-implementation period, from March to May 2019. After the implementation of the FOPWL law, a second wave of data collection occurred from March 2020 to February 2021. Although fieldworkers began data collection in person for the second wave during the first weeks of March 2020, the process was interrupted due to the COVID-19 pandemic restrictions. As a result, the missing food and beverage products were gradually purchased online to complete the dataset, a process that lasted up until February 2021. During both waves of data collection, photos were taken of all sides of food and beverage packages, either as they were available in-store or upon receipt from online purchases during the second wave.

In this study, the data collection and entry procedures were based on the photographic methods proposed by Kanter et al. which were previously used in similar studies in Chile, Colombia, and the study of top-selling products in Peru (22–26). Following each data collection wave, two research assistants reviewed the photos to ensure image clarity. Then, trained fieldworkers– nutritionists and food engineers–proceeded to enter label information separately for each product, including the barcode, brand, flavor, ingredient list, and manufacturer. They also recorded nutritional information: the amount of energy and nutrients such as protein, carbohydrates, total sugars, sodium, total fats, and fat subtypes. Although the FOPWL policy had been established, Peru does not mandate an NFP on packaged foods and beverages. As a result, many products had to be excluded due to missing NFP data. The available nutrient quantities were programmatically standardized to a content per 100 g or 100 mL of the product and subsequently reviewed by trained nutritionists. If applicable, instructions for product reconstitution (e.g., powdered milks, condensed juices, etc.) were recorded to estimate the amount of energy and nutrients of the product as consumed, since nutrient thresholds apply to the reconstituted version of these products. In these cases, the nutritional composition of the ingredients used in preparation was estimated using data from the USDA Food and Nutrient Database for Dietary Studies (27).

All data was collected and managed using REDCap electronic data capture tools hosted at University of North Carolina at Chapel Hill (28, 29).

### Food and beverages groups

Similar to previous analyses (15), the packaged foods and beverages were categorized into 15 mutually exclusive groups based on consumption-based and nutritional criteria, comprising 12 food groups and 3 beverage groups. Food groups included in the analysis were: bread and savory bakery; breakfast cereals and bars; dairy based products; candies, sweet confectionery, and processed fruits; ice creams; nuts and savory snacks; ready-to-eat meals; processed meats, fish, and seafood; savory sauces, spreads, and fats; sweet bakery; sweet spreads; and yogurt. The beverages groups for this analysis were: Sugar-sweetened beverages; milk and milk-based drinks; and milk substitutes and formulas. **S2 Table and S3 Table** provide examples of products included in each category, along with the number of products per group.

### Study Design

Two complementary analyses were conducted to assess changes in the nutritional composition of packaged foods in Peru. First, a repeated cross-sectional analysis was performed using all eligible products collected at T0 and T1. Then, a longitudinal analysis was conducted using only products that were present in both T0 and T1. While the cross-sectional analysis reflects changes in the total food supply, the longitudinal approach isolates industry responses by focusing on reformulated products, eliminating the influence of new product entries, product discontinuations, and potential sampling differences between waves. In both analyses, comparisons included the amount of nutrients of concerns and the proportion of products “high in” (i.e., foods and beverages with the amount of nutrients above the initial cutoffs) or containing trans fats. Together, these analyses provide a comprehensive evaluation of how the food industry has adapted to regulatory pressures.

### Definition of analytical sample

A total of 10,127 products were photographed during March-May 2019, March-September 2020, and December 2020-February 2021. To create a comprehensive post-implementation (T1) sample, data from 2020 and 2021 were combined, with only the most recent version retained when the same product was collected in both years.

**Fig 1** illustrates the inclusion and exclusion of products in both the cross-sectional and longitudinal analyses. Products were excluded from both T0 and T1 samples for the following reasons: absence of a nutrition facts label (∼25% of total products in both samples); falling outside the scope of the regulation (e.g., unprocessed and minimally processed foods, culinary ingredients); duplication (i.e., identical product and size for the same data collection wave); missing data on nutrients of concern (∼9% of products with an NFP in T0 and ∼5% in T1); presence of an NFP in a foreign language; or absence of essential information required for analysis (e.g., barcode). Additionally, to ensure data accuracy, the total energy content reported on the product label was cross-verified using the Atwater general factors, by calculating the sum of the energy contributions from macronutrients: 4 kcal/g for proteins and carbohydrates, and 9 kcal/g for fats (30). Products with irreparable errors, such as implausible nutrient amounts (i.e., >20% difference), were subsequently excluded.

**Fig 1.**
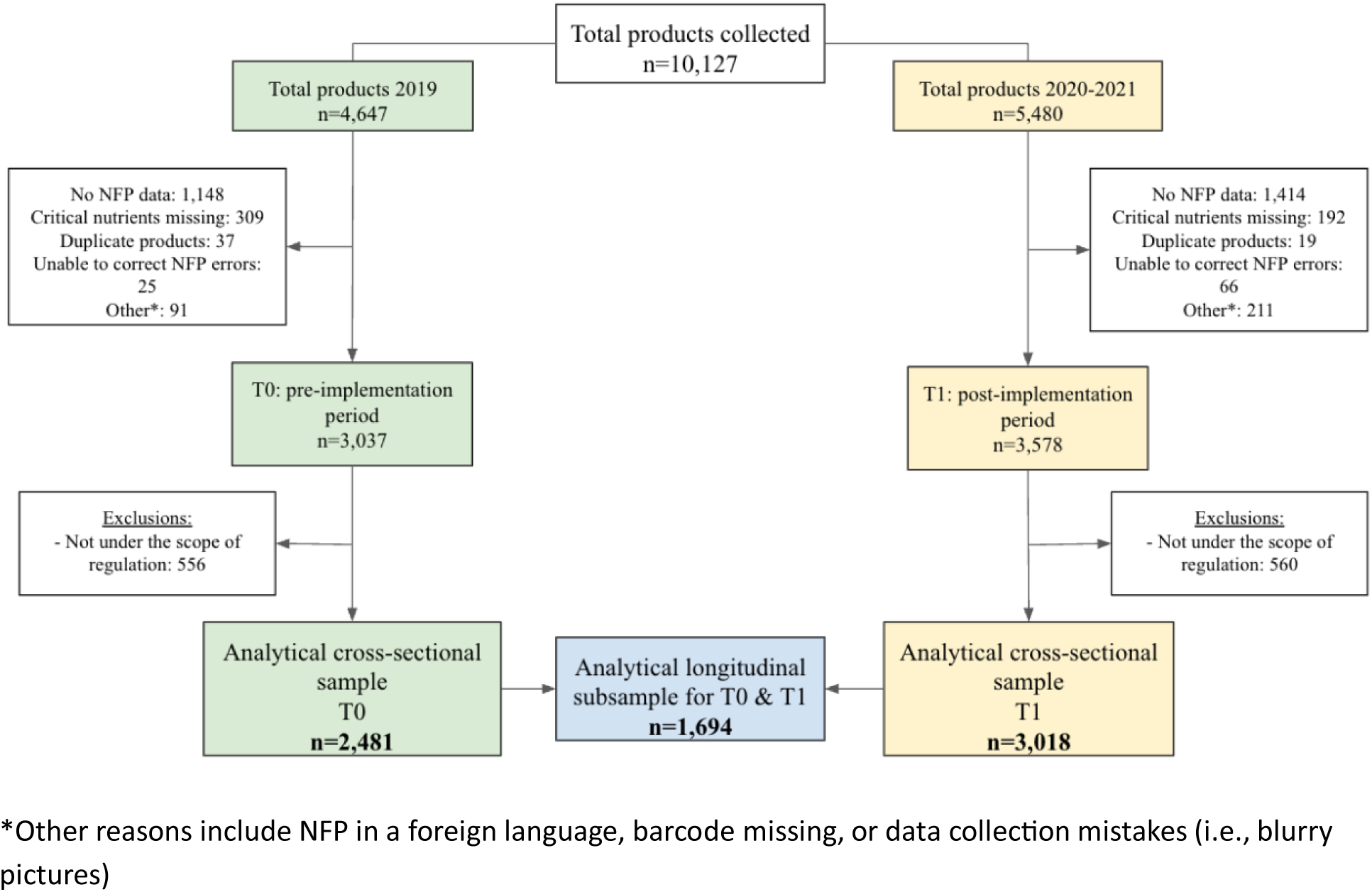
Flow diagram illustrating products included from the analytical dataset. T0, pre-implementation period; T1, post-implementation period; NFP, Nutrition Facts Panel. *Other reasons include NFP in a foreign language, barcode missing, or data collection mistakes (i.e., blurry pictures)

The final analytic sample for the repeated cross-sectional analysis included 2,481 products for the pre-implementation phase (T0) and 3,018 products for the post-implementation phase (T1). For the longitudinal analysis (i.e., products collected in both pre-and post-implementation periods), the sample included 1,694 products, representing approximately 68% of the T0 and 56% of the T1 items from the cross-sectional samples.

Products classified as “high in” nutrients were identified based on the criteria outlined in the first phase of the regulation. Nutritional cutoffs were applied to product as consumed according to the unit of measure specified on the label (i.e., grams for solids and milliliters for liquids). For the longitudinal analytical samples, products were programmatically matched across and within waves using barcode, brand, and product description.

## Data Analyses

### Study outcomes

The primary outcomes included (i) changes (i.e., reductions) in the proportion of products surpassing the cutoffs established in the first phase of the law for total sugars, sodium, saturated fats, or any “high in” (i.e., products high in at least one nutrient of concern), and products containing trans fats (i.e., any product that would receive a warning label according to the policy), and (ii) changes (i.e., reductions) in the quartiles of total sugars, sodium, and saturated fats (amount per 100 g or 100 mL). Changes in quartiles, rather than only median values, were evaluated as they provide a more detailed understanding of how nutrient content is distributed across products and whether reformulation was concentrated in a specific segment of the market (e.g., products with the highest nutrient levels) or occurred more broadly. These changes were evaluated overall, for foods and beverages, and by food or beverage group, comparing the pre-implementation period and post-implementation period of the Peruvian FOPWL law.

### Statistical analyses

Changes were analyzed using different statistical approaches for the repeated cross-sectional and longitudinal analyses. In the repeated cross-sectional analyses, changes in the proportion of “high in” foods or beverages between T0 and T1 were estimated using the estimated marginal means from unadjusted logistic regression models. To assess shifts in the distribution of nutrients of concern, we conducted quantile regression analyses, using the implementation period as the independent variable to estimate changes in the 25th, 50th, and 75th percentiles of each nutrient of concern within food or beverage group. Because the “contains trans fats” label applies to any amount of trans fats, quartile analysis was not conducted for this outcome.

For the longitudinal analyses, we used logistic regression models to examine changes in the proportion of “high in” products, clustering standard errors at the product level to account for repeated observations. To assess changes in nutrient distributions over time, we applied linear fixed-effects quantile regression models, which included food group-level fixed-effects, a fixed time-effect, and group-time interaction terms to measure group-level change over time. All analyses were conducted using STATA/SE version 18.0, with significance set at p < 0.05.

## Results

### Cross-sectional results

The cross-sectional analyses comparing 2020-2021 to 2019 revealed a marked decrease in the overall proportion of foods and beverages carrying any “high in” warnings following the implementation of the first phase of the law. The proportion of foods with at least one warning label declined by 6 percentage points, from 62% to 56%, while for beverages, it decreased by 11 percentage points, from 32% to 21% (**Table 1**). The extent of reduction varied by nutrient and product category. The proportion of products classified as “high in” total sugars decreased, with a relative reduction of 10% among foods and 36% among beverages (i.e., the percentage decrease relative to their 2019 levels). Additionally, pronounced relative reductions were observed in the proportion of foods labeled as “high in” sodium (19%) and those containing trans fat (41%). When considering foods and beverages together, an overall reduction of approximately 5.7 percentage points in the proportion of products carrying any “high in” warning was observed (**S4 Table**).

**Table 1.** Cross-sectional changes in the proportion of “high in” nutrients of concern (or any high in) before (T0) and after the first phase of Peru’s Law (T1) for overall foods and beverages.

|  | 2019 (T<sub>0</sub>)<br>% (95% CI) | 2020 (T<sub>1</sub>)<br>% (95% CI) | Difference (T0 vs T1) |  |  |
| --- | --- | --- | --- | --- | --- |
|  |  |  | Absolute<br>change | Relative<br>change (% of<br>T0) | p-value |
| **Foods** | n = 1,839 | n = 2,383 |  |  |  |
| Any “High in” | 62.3 (60.0, 64.5) | 56.0 (54.0, 58.0) | −6.3 | −10.1 | <0.01 |
| High in Sugars | 41.0 (38.7, 43.3) | 36.8 (34.8, 38.8) | −4.1 | −10.1 | <0.01 |
| High in Sodium | 10.4 (9.0, 11.7) | 8.4 (7.3, 9.5) | −2.0 | −18.9 | 0.03 |
| High in Saturated Fats | 38.7 (36.5, 40.9) | 36.9 (34.9, 38.9) | −1.8 | −4.6 | 0.24 |
| Contains Trans Fats | 8.6 (7.4, 9.9) | 5.1 (4.3, 6.1) | −3.6 | −41.3 | <0.01 |
| **Beverages** | n = 642 | n = 635 |  |  |  |
| Any “High in” | 32.2 (28.6, 35.9) | 21.4 (18.2, 24.6) | −10.8 | −33.6 | <0.01 |
| High in Sugars | 31.7 (28.0, 35.3) | 20.4 (17.2, 23.5) | −11.3 | −35.6 | <0.01 |
| High in Sodium | 0.9 (0.2, 1.7) | 1.0 (0.2, 1.7) | 0.0 | 2.1 | 0.97 |
| High in Saturated Fats | 0.6 (0.0, 1.2) | 0.6 (0.0, 1.3) | 0.0 | 2.1 | 0.98 |
| Contains Trans Fats | 0 | 0 | NA | NA | NA |

Values represent the sample size and the proportion of regulated products.
Cutoffs correspond to the limits on the amount of energy or nutrient of concern for the implementation of the first phase of the law (i.e., for solids, per 100g: 22.5 g of sugars, 6 g of saturated fats, 800 mg of sodium; for liquids, per 100 mL: 6 g of sugars, 3 g of saturated fats, 100 mg of sodium).
T0: pre-implementation period, March to May 2019 (n = 2,481); T1: post-implementation of the 1st phase of the law, March 2020 to February 2021 (n = 3,018).

**Table 2** presents changes in the proportion of products carrying “high in” warnings across various food and beverage categories. The most pronounced relative reductions, with a strong evidence of a difference, were observed in products “high in” total sugars, in the order of 42%, 37%, 21%, and 15% in sugar-sweetened beverages, breakfast cereals and bars, sweet spreads, and sweet bakery products, respectively. Within the sugar-sweetened beverage category, further analysis indicated that the decline in products “high in” sugar was largely driven by reductions among juices and nectars (**S5 Table**). Declines in “high in” saturated fats products were noted in ready-to-eat meals (82% relative reduction) and sweet bakery items (18% relative reduction).

**Table 2.** Cross-sectional changes in the proportion of “high in” nutrients of concern (or any high in) before (T0) and after the first phase of Peru’s Law (T1) by food and beverage group.

|  | 2019 (T<sub>0</sub>)<br>% (95% CI) | 2020 (T<sub>1</sub>)<br>% (95% CI) | Difference (T0 vs T1) |  | p-value |
| --- | --- | --- | --- | --- | --- |
|  |  |  | Absolute | Relative (% of T0) |  |
| Foods Groups |  |  |  |  |  |
| Bread and Savory Bakery | N = 146 | N = 178 |  |  |  |
| Any “High in” | 23.3 (16.4, 30.1) | 18.5 (12.8, 24.2) | -4.7 | -20.4 | 0.30 |
| High in Sugars | 0 | 0 | NA | NA | NA |
| High in Sodium | 11.0 (5.9, 16.0) | 10.1 (5.7, 14.5) | -0.8 | -7.7 | 0.81 |
| High in Saturated Fats | 13.7 (8.1, 19.3) | 8.6 (4.4, 12.7) | -5.1 | -37.4 | 0.15 |
| Contains Trans Fats | 4.1 (0.9, 7.3) | 2.2 (0.1, 4.4) | -1.9 | -45.3 | 0.35 |
| Breakfast Cereals and Bars | N = 159 | N = 205 |  |  |  |
| Any “High in” | 66.0 (58.7, 73.4) | 42.0 (35.2, 48.7) | -24.1 | -36.5 | <0.01 |
| High in Sugars | 63.9 (56.4, 71.4) | 40.7 (33.9, 47.5) | -23.2 | -36.3 | <0.01 |
| High in Sodium | 0 | 0 | NA | NA | NA |
| High in Saturated Fats | 5.7 (2.1, 9.3) | 3.6 (1.0, 6.1) | -2.1 | -37.2 | 0.35 |
| Contains Trans Fats | 10.1 (5.4, 14.7) | 7.8 (4.1, 11.5) | -2.3 | -22.4 | 0.46 |
| Dairy Based products* | N = 62 | N = 99 |  |  |  |
| Any “High in” | 88.7 (80.8, 96.5) | 96.0 (92.1, 99.8) | 7.2 | 8.2 | 0.11 |
| High in Sugars | 1.6 (-1.5, 4.7) | 2.2 (-0.8, 5.3) | 0.6 | 37.8 | 0.79 |
| High in Sodium | 21.3 (11.0, 31.6) | 20.2 (12.3, 28.1) | -1.1 | -5.2 | 0.87 |
| High in Saturated Fats | 83.9 (74.7, 92.1) | 96.0 (92.1, 99.8) | 12.1 | 14.4 | 0.02 |
| Contains Trans Fats | Excluded | Excluded | NA | NA | NA |
| Candies, Sweet Confectionery, and Processed Fruit | N = 493 | N = 575 |  |  |  |
| Any “High in” | 76.9 (73.2, 80.6) | 81.9 (78.8, 85.1) | 5.0 | 6.6 | 0.04 |
| High in Sugars | 75.1 (71.2, 78.9) | 78.5 (75.2, 82.0) | 3.5 | 4.7 | 0.18 |
| High in Sodium | 1.8 (0.6, 3.0) | 1.7 (0.7, 2.8) | -0.1 | -4.7 | 0.92 |
| High in Saturated Fats | 52.4 (48.0, 56.9) | 58.8 (54.8, 62.9) | 6.4 | 12.1 | **0.04** |
| Contains Trans Fats | 10.8 (8.0, 13.5) | 6.8 (4.7, 8.8) | -4.0 | -36.9 | **0.02** |
| **Ice creams** | N = 45 | N = 70 |  |  |  |
| Any “High in” | 75.6 (63.0, 88.1) | 65.7 (54.6, 76.8) | -9.8 | -13.0 | 0.25 |
| High in Sugars | 57.8 (43.3, 72.2) | 40.0 (28.5, 51.5) | -17.8 | -30.8 | 0.06 |
| High in Sodium | 0 | 0 | NA | NA | NA |
| High in Saturated Fats | 48.9 (34.3, 63.5) | 54.3 (42.6, 66.0) | 5.4 | 11.0 | 0.57 |
| Contains Trans Fats | 0 | 0 | NA | NA | NA |
| **Nuts and savory snacks** | N = 170 | N = 205 |  |  |  |
| Any “High in” | 64.7 (57.5, 71.9) | 54.6 (47.8, 61.4) | -10.1 | -15.6 | **0.04** |
| High in Sugars | 5.9 (2.4, 9.5) | 4.0 (1.3, 6.7) | -1.9 | -32.1 | 0.41 |
| High in Sodium | 13.5 (8.4, 18.7) | 15.6 (10.6, 20.6) | 2.1 | 15.4 | 0.57 |
| High in Saturated Fats | 54.1 (46.6, 61.6) | 44.3 (37.5, 51.2) | -9.8 | -18.1 | 0.06 |
| Contains Trans Fats | 0.6 (-0.6, 1.7) | 2.4 (0.3, 4.6) | 1.9 | 314.6 | 0.13 |
| **Ready-to-eat meals** | N = 89 | N = 141 |  |  |  |
| Any “High in” | 31.5 (21.8, 41.1) | 9.9 (5.0, 14.9) | -21.5 | -68.4 | **<0.01** |
| High in Sugars | 0 | 0 | NA | NA | NA |
| High in Sodium | 1.1 (-1.2, 3.3) | 2.1 (-0.3, 4.5) | 1.0 | 89.4 | 0.54 |
| High in Saturated Fats | 15.7 (8.2, 23.3) | 2.9 (0.1, 5.6) | -12.9 | -81.8 | **<0.01** |
| Contains Trans Fats | 20.2 (11.9, 28.6) | 5.0 (1.4, 8.5) | -15.3 | -75.5 | **<0.01** |
| **Processed meats, fish, and seafood** | N = 103 | N = 186 |  |  |  |
| Any “High in” | 44.7 (35.1, 54.3) | 43.0 (35.9, 50.1) | -1.6 | -3.7 | 0.79 |
| High in Sugars | 0 | 0 | NA | NA | NA |
| High in Sodium | 42.7 (33.2, 52.3) | 31.4 (24.7, 38.0) | -11.4 | -26.6 | 0.06 |
| High in Saturated Fats | 15.5 (8.5, 22.5) | 26.9 (20.5, 33.4) | 11.4 | 73.3 | **0.02** |
| Contains Trans Fats | 4.9 (0.7, 9.0) | 2.2 (0.1, 4.2) | -2.7 | -55.7 | 0.25 |
| **Savory sauces, spreads, and fats** | N = 179 | N = 189 |  |  |  |
| Any “High in” | 65.9 (59.0, 72.9) | 53.4 (46.3, 60.6) | -12.5 | -18.9 | **0.01** |
| High in Sugars | 12.0 (7.1, 16.9) | 13.9 (8.8, 18.9) | 1.9 | 16.0 | 0.60 |
| High in Sodium | 43.8 (36.5, 51.1) | 30.3 (23.7, 36.9) | -13.5 | -30.8 | **<0.01** |
| High in Saturated Fats | 31.8 (24.9, 38.7) | 22.9 (16.9, 28.9) | -8.9 | -28.1 | 0.06 |
| Contains Trans Fats | 9.5 (5.2, 13.8) | 6.9 (3.3, 10.5) | -2.6 | -27.6 | 0.36 |
| **Sweet bakery** | N = 210 | N = 293 |  |  |  |
| Any “High in” | 87.1 (82.6, 91.7) | 79.2 (74.5, 83.8) | -8.0 | -9.1 | **0.02** |
| High in Sugars | 79.1 (73.6, 84.7) | 67.5 (62.1, 72.8) | -11.7 | -14.7 | **<0.01** |
| High in Sodium | 0 | 0 | NA | NA | NA |
| High in Saturated Fats | 75.4 (69.5, 81.2) | 61.7 (56.1, 67.3) | -13.6 | -18.1 | **<0.01** |
| Contains Trans Fats | 19.5 (14.2, 24.9) | 10.2 (6.8, 13.7) | -9.3 | -47.6 | **<0.01** |
| **Sweet spreads** | N = 65 | N = 89 |  |  |  |
| Any “High in” | 80.0 (70.3, 89.7) | 60.7 (50.5, 70.8) | -19.3 | -24.2 | **<0.01** |
| High in Sugars | 81.4 (71.4, 91.3) | 64.5 (53.7, 75.2) | -16.9 | -20.8 | **0.02** |
| High in Sodium | 0 | 0 | NA | NA | NA |
| High in Saturated Fats | 18.2 (8.0, 28.4) | 13.9 (6.3, 21.6) | -4.3 | -23.4 | 0.51 |
| Contains Trans Fats | 1.5 (-1.5, 4.5) | 1.1 (-1.0, 3.3) | -0.4 | -27.0 | 0.83 |
| **Yogurt\*** | N = 118 | N = 153 |  |  |  |
| Any “High in” | 0.8 (-0.8, 2.5) | 5.9 (2.2, 9.6) | 5.0 | 594.1 | **0.02** |
| High in Sugars | 0.8 (-0.4, 6.6) | 3.9 (0.8, 7.0) | 3.1 | 362.7 | 0.08 |
| High in Sodium | 0 | 0 | NA | NA | NA |
| High in Saturated Fats | 0 | 0 | NA | NA | NA |
| Contains Trans Fats | Excluded | Excluded | NA | NA | NA |
| **Beverage Groups** |  |  |  |  |  |
| **Sugar-sweetened Beverages** | N = 442 | N = 398 |  |  |  |
| Any “High in” | 32.5 (28.2, 36.9) | 19.3 (15.4, 23.2) | –13.2 | –40.6 | **<0.01** |
| High in Sugars | 32.2 (27.8, 36.6) | 18.6 (14.8, 22.4) | –13.6 | –42.2 | **<0.01** |
| High in Sodium | 0.7 (0.0, 1.5) | 0.5 (0.0, 1.2) | –0.0 | –25.3 | 0.75 |
| High in Saturated Fats | 0 | 0 | NA | NA | NA |
| Contains Trans Fats | 0 | 0 | NA | NA | NA |
| **Milk and milk-based drinks** | N = 127 | N = 148 |  |  |  |
| Any “High in” | 36.2 (27.9, 44.6) | 27.7 (20.5, 34.9) | –8.5 | –23.5 | 0.13 |
| High in Sugars | 35.4 (27.1, 43.8) | 25.7 (18.6, 32.7) | –9.8 | –27.5 | 0.08 |
| High in Sodium | 0 | 0 | NA | NA | NA |
| High in Saturated Fats | 2.4 (–0.3, 5.0) | 2.8 (0.1, 5.5) | 0.4 | 16.7 | 0.84 |
| Contains Trans Fats | 0 | 0 | NA | NA | NA |
| **Milk substitutes and formulas** | N = 73 | N = 89 |  |  |  |
| Any “High in” | 23.3 (13.6, 33.0) | 20.2 (11.9, 28.6) | –3.1 | –13.2 | 0.63 |
| High in Sugars | 21.4 (11.8, 31.0) | 19.5 (11.2, 27.9) | –1.9 | –8.8 | 0.77 |
| High in Sodium | 0 | 0 | NA | NA | NA |
| High in Saturated Fats | 0 | 0 | NA | NA | NA |
| Contains Trans Fats | 0 | 0 | NA | NA | NA |

However, an increase in the proportion of products “high in” saturated fats was observed for dairy-based products; processed meats, fish, and seafood; and candies, sweet confectionery, and processed fruits. For trans fats, relative reductions were found in candies, sweet confectionery, and processed fruits (82%), ready-to-eat meals (76%), and sweet bakery (48%). In contrast, a decrease in the proportion of “high in” sodium products was only noted in savory sauces, spreads, and fats (31% relative reduction). Additionally, certain categories, such as yogurts and candies, sweet confectionery, and processed fruits, experienced an increase in the proportion of products carrying at least one warning label.

**Table 3** presents cross-sectional quantile regression results examining changes in distribution of total sugars, saturated fats, and sodium before and after the implementation of the first phase of Peru’s FOPWL law, disaggregated by food and beverage categories. The largest changes were observed in product groups where the regulatory thresholds were set below the 75th percentile of the pre-policy nutrient distribution. However, shifts in nutrient levels were also observed below the established cutoffs for some categories (See examples of shifts in the distribution of sugars, sodium, and saturated fats in **Fig 2**).

**Fig 2.**
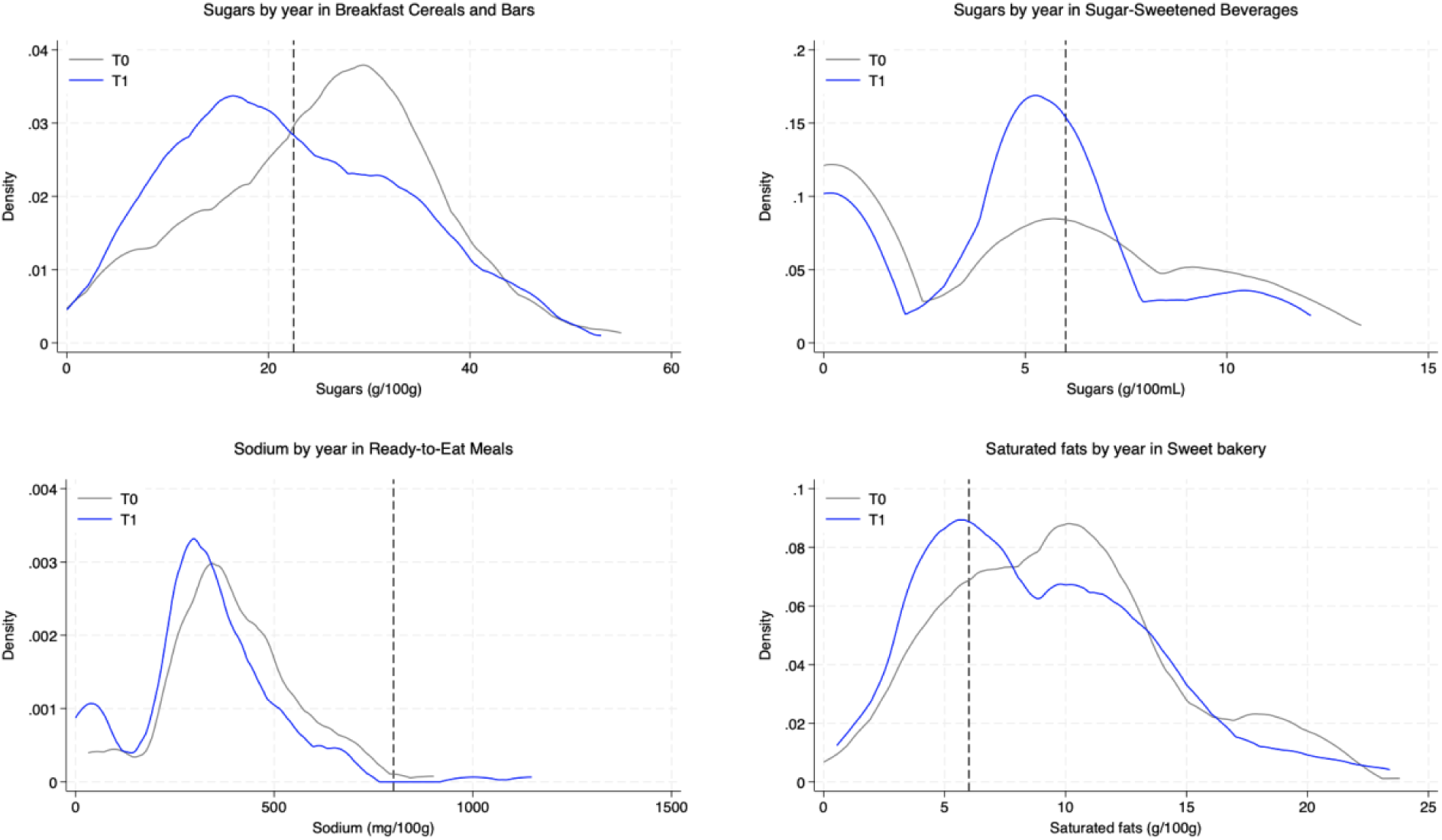
Density curves for the amount of total sugars in breakfast cereals and bars and sugar-sweetened beverages, sodium in ready-to-eat meals, and saturated fats in sweet bakery, repeated cross-sectional analysis. The gray line represents the distribution in T0 (preimplementation period, March-June 2019); the blue line represents distribution in T1 (postimplementation of the 1^st^ phase of the law, March 2020-February 2021); the black dashed line indicates the nutrient cutoff established for Phase 1 implementation of the law.

**Table 3.** Cross-sectional changes in quartiles of nutrients of concern before (T0) and after first phase of Peru’s Law (T1) for overall food and beverages and by food and beverage group.

|  | **TOTAL SUGARS**<br>**(g/100g-mL)** |  | **SODIUM**<br>**(mg/100g-mL)** |  | **SATURATED FATS**<br>**(g/100g-mL)** |  |
| --- | --- | --- | --- | --- | --- | --- |
|  | 2019 (T0) | 2020-21 (T1) | 2019 (T0) | 2020-21 (T1) | 2019 (T0) | 2020-21 (T1) |
| **Overall Foods** |  |  |  |  |  |  |
| p25 | 2.2 | 2.0 | 66.7 | 69.0 | 0.3 | 0.5 |
| p50 | 17.2 | 15.0 | 268.0 | 251.2 | 4.0 | 4.1 |
| p75 | 40.0 | 37.5 | 494.1 | 480.0 | 11.2 | 11.8 |
| **Overall Beverages** |  |  |  |  |  |  |
| p25 | 1.7 | **4.0** | 7.5 | **9.0** | 0 | 0 |
| p50 | 5.7 | **5.4** | 31.4 | 35.3 | 0 | 0 |
| p75 | 9.3 | 8.5 | 50.0 | 51.5 | 1.0 | 1.1 |
| **Food Groups** |  |  |  |  |  |  |
| **Bread and Savory Bakery** |  |  |  |  |  |  |
| T0 cutoff | NA |  | 90<sup>th</sup> percentile |  | 86<sup>th</sup> percentile |  |
| p25 | 3.0 | 2.8 | 339.0 | 366.2 | 0.9 | 1.0 |
| p50 | 4.0 | 4.7 | 456.3 | 498.6 | 2.0 | 2.0 |
| p75 | 6.8 | 7.3 | 706.7 | 718.8 | 3.8 | 4.6 |
| **Breakfast Cereals and Bars** |  |  |  |  |  |  |
| T0 cutoff | 37<sup>th</sup> percentile |  | NA |  | 96<sup>th</sup> percentile |  |
| p25 | 17.0 | 13.6 | 157.0 | **81.2** | 0 | 0 |
| p50 | 26.7 | **20.0** | 311.0 | 273.0 | 0.7 | **0.2** |
| p75 | 32.2 | 31.2 | 410.0 | 375.0 | 2.4 | 2.1 |
| **Dairy Based Products** |  |  |  |  |  |  |
| T0 cutoff | 99<sup>th</sup> percentile |  | 78<sup>th</sup> percentile |  | 11<sup>th</sup> percentile |  |
| p25 | 0.1 | 0 | 73.6 | 70.0 | 11.0 | 11.7 |
| p50 | 2.1 | 2.0 | 550.0 | 480.0 | 15.0 | 16.7 |
| p75 | 3.3 | 3.6 | 781.0 | 746.7 | 19.3 | 20.0 |
| **Candies, Sweet Confectionery, and Processed Fruit** |  |  |  |  |  |  |
| T0 cutoff | 25<sup>th</sup> percentile |  | 98<sup>th</sup> percentile |  | 47<sup>th</sup> percentile |  |
| p25 | 24.5 | 31.8 | 10.0 | **20.0** | 0 | 0 |
| p50 | 49.0 | 47.0 | 51.8 | 53.0 | 9.4 | 12.6 |
| p75 | 57.0 | 56.0 | 95.0 | 96.0 | 18.5 | 18.5 |
| Ice Cream |  |  |  |  |  |  |
| T0 cutoff | 42<sup>nd</sup> percentile |  | NA |  | 51<sup>st</sup> percentile |  |
| p25 | 21.2 | 20.5 | 59.0 | 58.0 | 3.6 | 3.5 |
| p50 | 23.2 | **21.8** | 72.0 | 74.7 | 6.0 | 6.6 |
| p75 | 24.0 | 25.3 | 85.0 | 86.0 | 9.6 | 9.5 |
| Nuts and Savory Snacks |  |  |  |  |  |  |
| T0 cutoff | 91<sup>st</sup> percentile |  | 86<sup>th</sup> percentile |  | 48<sup>th</sup> percentile |  |
| p25 | 0.4 | 0.4 | 320.0 | 279.0 | 3.5 | 2.8 |
| p50 | 3.3 | 2.9 | 440.0 | 422.2 | 6.7 | 5.4 |
| p75 | 6.7 | 6.0 | 660.0 | 710.0 | 10.0 | 10.0 |
| Ready-To-Eat Meals |  |  |  |  |  |  |
| T0 cutoff | NA |  | 99<sup>th</sup> percentile |  | 91<sup>st</sup> percentile |  |
| p25 | 0 | **0.3** | 301.0 | 258.5 | 0.5 | 0.3 |
| p50 | 0.5 | 0.6 | 390.5 | **314.5** | 1.3 | 1.2 |
| p75 | 1.5 | 2.0 | 483.7 | **405.8** | 2.6 | 1.9 |
| Processed Meats, Fish, and Seafood |  |  |  |  |  |  |
| T0 cutoff | NA |  | 57<sup>th</sup> percentile |  | 81<sup>st</sup> percentile |  |
| p25 | 0 | 0 | 400.0 | 393.0 | 0.8 | 1.0 |
| p50 | 0 | 0 | 677.4 | 600.0 | 2.0 | **3.5** |
| p75 | 0.5 | 0.8 | 1110.0 | 929.8 | 4.5 | 6.2 |
| Savory Sauces, Spreads, and Fats |  |  |  |  |  |  |
| T0 cutoff | 90<sup>th</sup> percentile |  | 56<sup>th</sup> percentile |  | 68<sup>th</sup> percentile |  |
| p25 | 0.0 | 0.0 | 480.0 | 415.0 | 0 | 0 |
| p50 | 2.7 | 4.0 | 721.4 | 678.6 | 1.0 | 0.4 |
| p75 | 7.0 | 7.1 | 1000.0 | 944.4 | 7.1 | **4.7** |
| Sweet Bakery |  |  |  |  |  |  |
| T0 cutoff | 21<sup>st</sup> percentile |  | NA |  | 26<sup>th</sup> percentile |  |
| p25 | 23.3 | 22.1 | 160.0 | 136.0 | 6.0 | 5.5 |
| p50 | 30.2 | 29.0 | 242.0 | **222.0** | 9.7 | **8.3** |
| p75 | 37.5 | 36.7 | 342.1 | 333.3 | 12.0 | 12.0 |
| Sweet Spreads |  |  |  |  |  |  |
| T0 cutoff | 19<sup>th</sup> percentile |  | NA |  | 82<sup>nd</sup> percentile |  |
| p25 | 33.2 | **18.8** | 15.0 | 13.8 | 0 | 0 |
| p50 | 47.0 | 47.0 | 44.0 | 31.5 | 0 | 0 |
| p75 | 59.0 | 59.0 | 125.0 | 103.0 | 5.0 | 4.5 |
| Yogurt |  |  |  |  |  |  |
| T0 cutoff | 99<sup>th</sup> percentile |  | NA |  | NA |  |
| p25 | 6.0 | **9.0** | 46.0 | 47.0 | 0 | 0.1 |
| p50 | 10.1 | 11.0 | 54.2 | **50.0** | 1.0 | 1.1 |
| p75 | 14.0 | 13.9 | 71.9 | 74.0 | 1.5 | 1.5 |
| Beverages Groups |  |  |  |  |  |  |
| Sugar-sweetened beverages |  |  |  |  |  |  |
| T0 cutoff | 70<sup>th</sup> percentile |  | 99<sup>th</sup> percentile |  | NA |  |
| p25 | 0 | 0.3 | 4.2 | 5.0 | 0 | 0 |
| p50 | 4.7 | 5.0 | 9.0 | 9.0 | 0 | 0 |
| p75 | 7.5 | **5.9** | 22.6 | 20.0 | 0 | 0 |
| Milk and Milk-Based Drinks |  |  |  |  |  |  |
| T0 cutoff | 59<sup>th</sup> percentile |  | 95<sup>th</sup> percentile |  | 96<sup>th</sup> percentile |  |
| p25 | 4.8 | 4.8 | 45.6 | 43.0 | 1.0 | 1.0 |
| p50 | 5.8 | **5.0** | 51.0 | 54.0 | 1.4 | 1.3 |
| p75 | 9.0 | **6.1** | 57.0 | **63.0** | 2.0 | 2.0 |
| Milk Substitutes and Formulas |  |  |  |  |  |  |
| T0 cutoff | 61<sup>st</sup> percentile |  | 96<sup>th</sup> percentile |  | 99<sup>th</sup> percentile |  |
| p25 | 2.8 | 2.9 | 36.5 | 35.2 | 0.2 | 0.2 |
| p50 | 5.0 | 4.9 | 40.0 | 40.0 | 0.7 | 0.7 |
| p75 | 6.7 | 5.8 | 50.0 | 55.0 | 1.0 | 1.1 |

T0: pre-implementation period, March to May 2019 (n = 2,481); T1: post-implementation of the 1st phase of the law, March 2020 to February 2021 (n = 3,018).
Quartiles and p-values were obtained from quantile regression models (one model per nutrient per food or beverage group), using the implementation period as independent variable. Significant p-values are bold and represent a p-value <0.05 versus T0.

For total sugars, no major shifts were detected in the overall distribution among foods. However, a reduction was observed in overall beverages at the median, where sugar content declined from 5.7 to 5.4 g/100 mL. Among individual categories, sugar-sweetened beverages exhibited a pronounced decline at the 75th percentile, decreasing from 7.5 to 5.9 g/100 mL, a level that aligns closely with the regulatory sugar threshold of 6g per 100mL. Breakfast cereals and bars showed a reduction at the median, from 26.7 to 20.0 g/100 g, while sweet spreads declined at the 25th percentile, from 33.2 to 18.8 g/100 g. Sweet bakery products also showed a marginal reduction at the 25th percentile (from 23.3 to 22.1 g/100 g). Interestingly, several groups that did not experience changes in the proportion of items labeled “high in” sugar still demonstrated notable reductions at specific points in the distribution. For instance, milk and milk-based drinks experienced a reduction in sugar content at the 75th percentile (from 9.0 to 6.1 g/100 mL), while ice creams showed a decline at the median (from 23.2 to 21.8 g/100 g). In contrast, some food groups—namely, ready-to-eat meals, yogurt, and savory sauces, spreads, and fats—exhibited slight increases in sugar content at lower percentiles; however, these values remained below the regulatory thresholds for FOPWL.

Regarding sodium, no major reductions were observed in the sodium content of total foods or beverages. At the category level, modest reductions were noted in breakfast cereals and bars, ready-to-eat meals, sweet bakery products, and yogurt. In contrast, slight increases in sodium content were found in candies, sweet confectionery, processed fruit, and milk and milk-based drinks. Importantly, all of these changes occurred at levels well below the regulatory sodium threshold and thus did not affect FOPWL eligibility. Among savory sauces, spreads, and fats— the only group that exhibited a decrease in the proportion of products labeled “high in” sodium—there was weak evidence of a reduction near the regulatory cutoff at the 75th percentile, declining from 1000.0 to 944.4 mg/100 g.

For saturated fats, reductions were observed in select food categories. In sweet bakery products, the median saturated fat content decreased from 9.7 to 8.3 g/100 g. Similarly, savory sauces, spreads, and fats demonstrated a reduction at the 75th percentile, declining from 7.1 to 4.7 g/100g. Conversely, an increase in saturated fat content was detected among processed meats, fish, and seafood at the median level. However, this increase remained within the non–“high in” range and therefore did not affect the eligibility of these products for FOPWL.

### Longitudinal Results

**Table 4** presents the findings from the longitudinal analyses, revealing a major decrease in the proportion of foods carrying any “high in” warnings following the initial implementation of the law. The proportion of foods with at least one warning decreased by 6 percentage points, while the proportion of beverages dropped by 12 percentage points. Relative reductions observed in foods ranged from 26% in products “high in” trans fats to 7% among those containing saturated fats. Likewise, the proportion of beverages “high in” total sugars showed a substantial decline in the post-implementation period (33% relative reduction). When considering foods and beverages together, an overall reduction of approximately 7.5 percentage points in the proportion of products carrying any “high in” warning was observed (**S6 Table**).

**Table 4.** Longitudinal changes in the proportion of “high in” nutrients of concern (or any high in) before (T0) and after the first phase of Peru’s Law (T1) for overall foods and beverages.

|  | 2019 (T0)<br>% (95% CI) | 2020 (T1)<br>% (95% CI) | Difference (T0 vs T1) |  |  |
| --- | --- | --- | --- | --- | --- |
|  |  |  | Absolute<br>change | Relative change<br>(% of T0) | p-value |
| **Foods** | N = 1,279 |  |  |  |  |
| Any “High in” | 61.0 (58.3, 63.7) | 54.9 (52.2, 57.6) | −6.1 | −10.0 | <**0.01** |
| High in Sugars | 41.1 (38.4, 43.9) | 37.1 (34.4, 39.8) | −4.0 | −9.8 | <**0.01** |
| High in Sodium | 9.9 (8.2, 11.5) | 8.5 (7.0, 10.1) | −1.3 | −13.5 | <**0.01** |
| High in Saturated Fats | 37.0 (34.4, 40.0) | 34.3 (31.7, 36.9) | −2.8 | −7.4 | <**0.01** |
| Contains Trans Fats | 8.4 (6.9, 10.0) | 6.3 (4.9, 7.6) | −2.2 | −25.9 | <**0.01** |
| **Overall Beverages** | N = 415 |  |  |  |  |
| Any “High in” | 36.9 (32.2, 41.5) | 25.1 (20.9, 29.2) | −11.8 | −32.0 | <**0.01** |
| High in Sugars | 36.3 (31.7, 41.0) | 24.2 (20.1, 28.3) | −12.1 | −33.3 | <**0.01** |
| High in Sodium | 0.9 (0.0, 1.9) | 0.9 (0.0, 1.9) | 0.0 | 0.2 | 0.37 |
| High in Saturated Fats | 0.7 (0.0, 1.5) | 0.7 (0.0, 1.5) | 0.0 | 0.2 | 0.39 |
| Contains Trans Fats | 0 | 0 | NA | NA | NA |

Values represent the sample size and the proportion of regulated products.
Cutoffs correspond to the limits on the amount of energy or nutrient of concern for the implementation of the first phase of the law (i.e., for solids, per 100g: 22.5 g of sugars, 6 g of saturated fats, 800 mg of sodium; for liquids, per 100 mL: 6 g of sugars, 3 g of saturated fats, 100 mg of sodium).
T0: pre-implementation period, March to May 2019 (n = 1,694); T1: post-implementation of the 1st phase of the law, March 2020 to February 2021 (n = 1,694).

While longitudinal changes in food and beverage groups generally mirrored those observed in the cross-sectional analyses, some differences emerged (**S7 Table and S8 Table**). In addition to the food groups with declines in “high in” sugars seen in the cross-sectional analysis, the longitudinal analysis also showed relative reductions in the order of 33% and 24% in ice creams and milk and milk-based drinks, respectively. Similarly, additional relative reductions were observed in products “high in” saturated fats, ranging from 71% in breakfast cereals and bars to 50% in bread and savory bakery. In contrast, while the cross-sectional analysis identified reductions in trans fat-containing products across three food groups, the longitudinal analysis found a decrease only in sweet bakery products (56% relative reduction).

Similarly, the longitudinal quantile regression analyses largely mirrored the patterns observed in the cross-sectional models but also revealed nuances not captured in the cross-sectional approach (**S9 Table**). Notably, reductions in sugar content were detected in candies, sweet confectionery, and processed fruits, as well as in the sodium content of processed meats, fish, and seafood, despite no corresponding decrease in the proportion of products exceeding the “high in” thresholds. Additionally, while reductions in saturated fat were observed at the 25th percentile of sweet bakery products, a rightward shift was found at the same percentile for saturated fat in bread and savory bakery. Nevertheless, this increase remained well below the regulatory threshold and did not affect the overall “high in” classification.

## Discussion

This study examined changes in the amount of nutrients of concern in a large sample of packaged foods and beverages available in Peruvian markets following the first phase of the FOPWL policy. Our findings reveal that, in the cross-sectional analysis, the proportion of foods with any “high in” decreased by 6 percentage points, while the proportion of beverages with any “high in” decreased by 11 percentage points compared to the pre-implementation period.

Overall, the main reductions were observed in products “high in” sugars, with relative decreases of a 10% among foods items and 36% among beverages. Notable relative reductions, in the order of 40% and 19% were also observed in trans fats and sodium, respectively. The reductions varied by food and beverage categories, with the greatest changes occurring in groups where the regulatory thresholds were set below the 75th percentile of the nutrient distribution–suggesting that the policy incentivized reformulation among products with initially higher nutrient levels.

The longitudinal analysis confirmed these trends, reinforcing the finding that Peru’s FOPWL policy has influenced reformulation efforts. These findings suggest that nutrient warning labels can be an effective regulatory tool to encourage manufacturers to improve the nutritional quality of packaged foods, potentially contributing to healthier food environments and reducing diet-related health risks.

In this section, we interpret our main findings in the context of previous studies from Peru and other Latin American countries, compare cross-sectional and longitudinal analyses, explore possible mechanisms driving observed changes, discuss implications for policy and public health, and acknowledge study limitations.

Peru was the second country to implement a FOPWL law in Latin America and the first middle-income country in the region to do so. Limited evidence existed on how Peru’s FOPWL policy has influenced the overall nutritional quality of the food supply. Our study expands upon an initial study that examined changes two years after the policy implementation in nutrients of concern in the top-selling products in Peru, which found a decline in the proportion of “high in” foods and beverages after the law’s implementation (23). Both studies found reductions in sugar content and in the proportion of products “high in” sugars. Additionally, both studies observed a decrease in the proportion of foods “containing trans fats.” However, while Saavedra-Garcia et al. (23) found no reduction in the proportion of foods “high in” sodium or saturated fats, our analyses yielded different findings. Specifically, our cross-sectional and longitudinal analyses revealed a reduction in the proportion of foods “high in” sodium, while the longitudinal analyses also identified a decrease in the proportion of foods “high in” saturated fats. These differences may be attributed to variations in product selection and sample size; the earlier study included only 94 longitudinal products, while ours evaluated over 1,600 products across multiple food and beverage categories.

In addition, our study methodology and results align with findings from Chile following the first phase of its FOPWL implementation. Cross-sectional analyses indicate that the magnitude of change was comparable in both countries: in Chile, the proportion of products (i.e., both foods and beverages) with any “high in” designation decreased from 51% to 44% (a 7 percentage-point reduction) (15), while in Peru, it declined from 55% to 49% (approximately 6 percentage points). Both countries implemented their FOPWL policies approximately one year after increasing taxes on high-sugar beverages, which may have prompted reformulation of these products prior to the FOPWL rollout, though this was not assessed in our study. It is also important to note key differences between the two policies. While Chile’s regulations require a “high in energy” label, the Peruvian law does not. Conversely, Peru mandates a “contains trans fat” label, which is not required in Chile. Despite these differences, both countries saw the most pronounced reductions in products labeled “high in” sugars (15). However, while Chile experienced a reduction in the proportion of all products “high in” sodium, in Peru, this reduction was observed only among food products. This may reflect Peru’s already high compliance with the Pan American Health Organization’s regional sodium targets, both before and after the FOPWL policy’s implementation (31, 32). Given that further reductions were observed in Chile as nutrient thresholds became stricter in later phases of implementation (16), similar trends may be expected in Peru as its policy tightened in the second phase, implemented in September 2021. Overall, these results underscore the potential of mandatory labeling policies to drive meaningful product reformulation and enhance the healthfulness of the food supply (14, 33). While voluntary systems such as the Health Star Rating have also been linked to reformulation (12), mandatory policies are expected to produce broader and more consistent changes across the market (34).

The cross-sectional analyses captured changes in the prevalence of “high in” products and nutrients of concern in the food supply, reflecting not only reformulation but also changes due to the introduction of new products and the discontinuation of existing ones. In contrast, the longitudinal analyses provided stronger evidence of reformulation, as they tracked modifications in the same products over time. While both approaches yielded largely consistent results, key differences highlight their complementarity in assessing food supply changes. Both analyses identified reductions in sugar across breakfast cereals and bars, sweet bakery products, sweet spreads, and SSBs, as well as reductions in sodium in savory sauces and spreads, and in saturated fat in ready-to-eat meals and sweet bakery products, suggesting substantial reformulation in these categories. However, reductions in sugar, in ice cream and milk-based drinks, and saturated fat, in bread and savory bakery products and breakfast cereals and bars, were observed only in the longitudinal analysis, suggesting that new high-sugar and high-saturated fat products may have entered the market for these food and beverage groups, offsetting overall reductions in the cross-sectional sample. Conversely, both analyses identified increased saturated fat in dairy-based products, possibly due to reformulation choices favoring full-fat ingredients due to consumer preferences or industry trends. Increases in saturated fat in candies, sweet confectionery, processed fruits, and processed meats were observed only in the cross-sectional analysis, indicating that product turnover rather than direct reformulation may have driven these changes.

Additionally, trans fat reductions were evident in three food groups in the cross-sectional analysis (i.e., sweet bakery; candies, sweet confectionery, and processed fruits; and ready-to-eat meals), while the longitudinal analysis detected reductions only in sweet bakery. This is reflected in the greater relative decline in trans fat-containing products in the cross-sectional sample (40%) compared to the longitudinal sample (24%). These findings may reflect the effects of the regional and country regulatory efforts. Peru is one of the few countries that implemented a specific label indicating “contains trans fats.” Important efforts have been made to reduce and eliminate the presence of trans fats in the food supply, including a Supreme Decree issued in 2016 that mandated their gradual elimination from the food supply (35, 36). Our findings suggest that manufacturers have increasingly phased out products containing trans fats or introduced new technologies and formulations of products not containing it, leading to a higher proportion of trans fat-free products in the food supply. Although some evidence indicates that removing trans fats may result in higher saturated fat content (37), this substitution effect was not observed in our analysis.

Furthermore, as previously noted, the most substantial reductions in both cross-sectional and longitudinal analyses were observed in the proportion of products classified as “high in” total sugars. A key factor contributing to this substantial decline may be the food industry’s ability to maintain sweetness levels while reducing sugar content by incorporating non-nutritive sweeteners (NNS) to avoid the FOPWL (38). Unlike policies in Mexico and Argentina, Peru’s FOPWL law does not require labels for NNS, potentially incentivizing manufacturers to replace sugar with these alternative sweeteners (17). Evidence from Chile supports this concern, as the use of NNS increased following the implementation of its labeling policy, with reformulated products that lowered sugar content being more likely to incorporate NNS (39). A similar trend was observed in Peru’s top-selling products, where the proportion containing NNS increased from 34.5% in 2019 to 62.1% in 2021 (23). In contrast, Mexico’s recent evaluation found that the proportion of products containing NNS actually decreased, highlighting the effects of mandatory NNS labels (17). Further research is needed to assess shifts in the use of NNS and other additives across the broader Peruvian food supply and the potential health implications of increased consumption.

An important issue arises regarding the regulation of sugar content in yogurts. While both cross-sectional and longitudinal quantile regression analyses confirm that this food group contains elevated amounts of sugar, the proportion of items labeled as “high in” sugar is lower than expected (e.g., only 4% carried the label in the post-policy cross-sectional sample). This discrepancy stems from how these products report their unit of measurement. According to Peruvian regulations, semi-solid foods can be labeled either in grams or milliliters, at the discretion of the manufacturer. Most producers declare them in grams, which subjects these products to the more lenient nutrient thresholds applicable to solids (e.g., 22.5 g of sugar per 100 g instead of 6 g per 100 mL for liquids). As a result, many yogurts that would otherwise qualify for FOPWL under a stricter liquid-based threshold avoid the label. This regulatory gap highlights a critical limitation in the policy’s design, as it may enable manufacturers to strategically classify products to evade stricter labeling requirements. Future revisions of the law should address this inconsistency within this food group.

Reformulation has been recognized as a key strategy for obesity prevention (33). In line with Geoffrey Rose’s concept of population-level prevention (40), our findings suggest that structural interventions like FOPWL can promote product reformulation that shifts the distribution of nutrients of concern in the food supply in a favorable direction (Fig 2). While the absolute reductions in “high in” product prevalence may appear modest, ranging from 6% to 11%, these changes are substantial in the context of public health. When these leftward shifts in nutrients of concern occur across a wide range of products consumed by millions, they may significantly reduce population-level exposure to diet-related risk factors for NCDs. Importantly, reformulation does not rely on individual behavior change and may help reduce health disparities by benefiting groups that face structural barriers to accessing healthier foods (41).

However, while reformulation plays a role in public health strategies, its limitations must also be acknowledged. Reformulated products may still be highly processed and contain alternative ingredients, such as artificial additives, which do not necessarily contribute to a healthier diet (42). Evidence from qualitative studies also suggests that consumers may perceive UPFs and SSBs without warning labels as inherently healthy, which could lead to unintended consequences (43). Moreover, there is limited long-term evidence on the extent to which reformulation translates into meaningful improvements in dietary intake and health outcomes. Thus, while reformulation may yield valuable public health outcomes, a comprehensive food policy framework is needed–one that both supports reformulation efforts to reduce harmful nutrients in processed foods and ensures equitable access to fresh, minimally processed options (44).

This study is not without limitations. First, its cross-sectional design does not allow us to disentangle the effects of reformulation, product entry, and product exit. However, our longitudinal analysis of products available before and after implementation provides additional evidence that reformulation played a key role in the observed changes. Second, because the inclusion of an NFP is not mandatory in Peru, we excluded a substantial proportion of products that lacked an NFP or had missing values for undeclared nutrients (∼34% of products at T0 and ∼30% at T1). It is possible that manufacturers of products high in nutrients of concern are more likely to omit NFPs to avoid disclosing unfavorable nutrient content. As a result, the products included in our analysis may be skewed toward relatively healthier options. This limitation reduced our ability to assess a more comprehensive sample of the food supply and underscores the need for policies mandating NFP inclusion on all packaged products in Peru. Third, we did not conduct laboratory analyses; instead, we relied on nutrient content information reported on product labels, which may not always reflect actual nutrient levels (45). Fourth, the study focused only on nutrients of concern, without evaluating whether these were replaced with other components such as additives, which was beyond the study’s scope. Finally, we did not assess the impact of the second phase of the FOPWL policy, which introduced stricter thresholds and may have led to further reformulation. Our findings reflect only the first phase and may underestimate longer-term changes.

Despite these limitations, this study provides valuable insights into the early effects of Peru’s FOPWL policy. The observed reductions in nutrients of concern suggest that the policy is driving positive changes in the food supply. However, continued monitoring is essential to assess long-term compliance and the impact of stricter regulatory thresholds. Notably, small-sized products—often more affordable and popular among children—are currently exempt from displaying warning labels despite exceeding nutrient thresholds (46). Future research should explore whether manufacturers exploit this exemption by maintaining less healthy formulations in smaller product sizes. Given that the Peruvian FOPWL is now fully implemented, further research should assess whether stricter thresholds in the second phase of implementation led to additional reformulation. Additionally, evaluating the broader dietary and health impacts of these changes, including shifts in consumer behavior and product substitution patterns, will be critical in shaping future regulatory strategies and informing food policies in other countries considering similar measures.

## Conclusion

In summary, our study demonstrates that the first phase of Peru’s FOPWL policy is associated with marked reductions in nutrients of concern in packaged foods and beverages. Both cross-sectional and longitudinal analyses revealed a substantial decrease in the proportion of products classified as “high in” sugars, sodium, saturated fats, and contains trans fats, underscoring the policy’s capacity to drive reformulation across diverse food and beverage categories. The greatest improvements were observed in categories where baseline nutrient levels were closer to the regulatory thresholds, suggesting that manufacturers are strategically reformulating products to avoid mandatory warnings. These findings parallel reformulation trends observed in other Latin American countries, yet also underscore unique differences in policy impact, particularly with respect to nutrients such as trans fats, which have only been regulated in Peru. The observed leftward shifts in the distribution of nutrients of concern reflect the type of population-level change envisioned in public health prevention strategies. This study contributes to the growing evidence of the public health potential of mandatory FOPWL policies. Ongoing monitoring is needed to assess the long-term compliance and health impacts as Peru implements stricter thresholds in subsequent policy phases, and future research should evaluate how these nutritional improvements translate into meaningful changes in dietary intake and health outcomes at the population level.

## Data Availability

All data produced in the present study are available upon reasonable request to the authors.

## Acknowledgements

We thank Bloomberg Philanthropies for financial support. Language editing assistance for select subsections of the Methods and Results was obtained using ChatGPT (OpenAI, version GPT-4) under the authors’ supervision to simplify and refine phrasing. None of the authors have conflict of interests of any type with respect to this manuscript.

## Supporting Information

## References

1. World Obesity Federation. World Obesity Atlas 2024. London: World Obesity Federation, 2024.

2. Instituto Nacional de Estadística e Informática. Perú: Enfermedades no Transmisibles y Transmisibles, 2020. 2020.

3. Instituto Nacional de Estadística e Informática. Perú: Enfermedades No Transmisibles y Transmisibles 2023. Lima, Perú: 2024.

4. Organización Panamericana de la Salud. Alimentos y bebidas ultraprocesados en América Latina: ventas, fuentes, perfiles de nutrientes e implicaciones. Washington, D.C.: 2019.

5. Meza-Hernández M, Villarreal-Zegarra D, Saavedra-Garcia L. Nutritional Quality of Food and Beverages Offered in Supermarkets of Lima According to the Peruvian Law of Healthy Eating. Nutrients. 2020;12(5):1508. doi: 10.3390/nu12051508.

6. Popkin BM, Reardon T. Obesity and the food system transformation in <scp>Latin America</SCP>. Obesity Reviews. 2018;19(8):1028–64. doi: 10.1111/obr.12694.

7. Hall KD, Ayuketah A, Brychta R, Cai H, Cassimatis T, Chen KY, et al. Ultra-Processed Diets Cause Excess Calorie Intake and Weight Gain: An Inpatient Randomized Controlled Trial of <EM>Ad Libitum</EM> Food Intake. Cell Metabolism. 2019;30(1):67–77.e3. doi: 10.1016/j.cmet.2019.05.008.

8. Pagliai G, Dinu M, Madarena MP, Bonaccio M, Iacoviello L, Sofi F. Consumption of ultra-processed foods and health status: a systematic review and meta-analysis. British Journal of Nutrition. 2021;125(3):308–18. doi: 10.1017/s0007114520002688.

9. Monteiro CA, Cannon G, Lawrence M, Louzada MLdC, Machado PP. Ultra-processed foods, diet quality, and health using the NOVA classification system. Rome: FAO, 2019.

10. Ley de Promoción de la Alimentación Saludable para Niños, Niñas y Adolescentes, Stat. LEY No 30021 (17 de Mayo, 2013, 2013).

11. Manual de Advertencias Publicitarias en el marco de lo establecido en la Ley N° 30021, Ley de promoción de la alimentación saludable para niños, niñas y adolescentes, y su Reglamento aprobado por Decreto Supremo N° 017-2017-SA, Stat. DECRETO SUPREMO N° 012-2018-SA (2018).

12. Roberto CA, Ng SW, Ganderats-Fuentes M, Hammond D, Barquera S, Jauregui A, et al. The Influence of Front-of-Package Nutrition Labeling on Consumer Behavior and Product Reformulation. Annual Review of Nutrition. 2021;41(1):529–50. doi: 10.1146/annurev-nutr-111120-094932.

13. Ganderats-Fuentes M, Morgan S. Front-of-Package Nutrition Labeling and Its Impact on Food Industry Practices: A Systematic Review of the Evidence. Nutrients. 2023;15(11):2630. doi: 10.3390/nu15112630.

14. Taillie LS, Duran AC. The case for mandatory - not voluntary - front-of-package nutrition labels. Bull World Health Organ. 2024;102(10):765–8. Epub 20240911. doi: 10.2471/blt.24.292537. PubMed PMID: 39318887; PubMed Central PMCID: PMC11418843.

15. Reyes M, Smith Taillie L, Popkin B, Kanter R, Vandevijvere S, Corvalán C. Changes in the amount of nutrient of packaged foods and beverages after the initial implementation of the Chilean Law of Food Labelling and Advertising: A nonexperimental prospective study. PLOS Medicine. 2020;17(7):e1003220. doi: 10.1371/journal.pmed.1003220.

16. Rebolledo N, Ferrer-Rosende P, Reyes M, Smith Taillie L, Corvalán C. Changes in the critical nutrient content of packaged foods and beverages after the full implementation of the Chilean Food Labelling and Advertising Law: a repeated cross-sectional study. BMC Medicine. 2025;23(1). doi: 10.1186/s12916-025-03878-6.

17. Salgado JC, Pedraza LS, Contreras-Manzano A, Aburto TC, Tolentino-Mayo L, Barquera S. Product reformulation in non-alcoholic beverages and foods after the implementation of front-of-pack warning labels in Mexico. PLOS Medicine. 2025;22(3):e1004533. doi: 10.1371/journal.pmed.1004533.

18. Reglamento que Establece el Proceso de Reducción Gradual Hasta la Eliminación de las Grasas Trans en Alimentos y Bebidas no Alcohólicas Procesados Industrialmente, (2016).

19. Decreto Supremo que modifica el Impuesto Selectivo al Consumo aplicable a los bienes del Nuevo Apéndice IV del TUO de la Ley del Impuesto General a las Ventas e Impuesto Selectivo al Consumo y el Reglamento de la Ley del Impuesto a la Renta, (2019).

20. Ministry of Health Peru. Lineamientos Para La Promoción Y Protección De La Alimentación Saludable En Las Instituciones Educativas Públicas Y Privadas De La Educación Básica. Lima, Peru 2019.

21. Diez-Canseco F, Cavero V, Alvarez Cano J, Saavedra-Garcia L, Taillie LS, Dillman Carpentier FR, et al. Design and approval of the nutritional warnings’ policy in Peru: Milestones, key stakeholders, and policy drivers for its approval. PLOS Global Public Health. 2023;3(6):e0001121. doi: 10.1371/journal.pgph.0001121.

22. Kanter R, Reyes M, Corvalán C. Photographic methods for measuring packaged food and beverage products in supermarkets. Current developments in nutrition. 2017;1(10):e001016.

23. Saavedra-Garcia L, Meza-Hernández M, Diez-Canseco F, Taillie LS. Reformulation of Top-Selling Processed and Ultra-Processed Foods and Beverages in the Peruvian Food Supply after Front-of-Package Warning Label Policy. International Journal of Environmental Research and Public Health. 2022;20(1):424. doi: 10.3390/ijerph20010424.

24. Taillie LS, Reyes M, Colchero MA, Popkin B, Corvalán C. An evaluation of Chile’s Law of Food Labeling and Advertising on sugar-sweetened beverage purchases from 2015 to 2017: A before-and-after study. PLoS medicine. 2020;17(2):e1003015.

25. Lowery CM, Mora-Plazas M, Gómez LF, Popkin B, Taillie LS. Reformulation of packaged foods and beverages in the Colombian food supply. Nutrients. 2020;12(11):3260.

26. Forero LC, Gómez LF, Mora-Plazas M, Parra-Murillo M, Toquica S, Taillie LS. Changes in the nutrient composition of top-selling packaged foods and beverages in Colombia between 2016 to 2021. Frontiers in Nutrition. 2025;12:1534195.

27. U.S. Department of Agriculture ARS. USDA Food and Nutrient Database for Dietary Studies 2017-2018. USDA, 2020.

28. Harris PA, Taylor R, Thielke R, Payne J, Gonzalez N, Conde JG. Research electronic data capture (REDCap)—A metadata-driven methodology and workflow process for providing translational research informatics support. Journal of Biomedical Informatics. 2009;42(2):377–81. doi: 10.1016/j.jbi.2008.08.010.

29. Harris PA, Taylor R, Minor BL, Elliott V, Fernandez M, O’Neal L, et al. The REDCap consortium: Building an international community of software platform partners. Journal of Biomedical Informatics. 2019;95:103208. doi: 10.1016/j.jbi.2019.103208.

30. Merrill AL, Watt BK. Energy value of foods: basis and derivation: Human Nutrition Research Branch, Agricultural Research Service, US…; 1955.

31. Blanco-Metzler A, Vega-Solano J, Franco-Arellano B, Allemandi L, Larroza RB, Saavedra-Garcia L, et al. Changes in the Sodium Content of Foods Sold in Four Latin American Countries: 2015 to 2018. Nutrients. 2021;13(11):4108. doi: 10.3390/nu13114108.

32. Yang Y, Flexner N, Tiscornia MV, Guarnieri L, Blanco-Metzler A, Núñez-Rivas H, et al. Monitoring sodium content in packaged foods sold in the Americas and compliance with the updated regional sodium reduction targets. PLOS ONE. 2025;20(4):e0304922. doi: 10.1371/journal.pone.0304922.

33. Gressier M, Swinburn B, Frost G, Segal AB, Sassi F. What is the impact of food reformulation on individuals’ behaviour, nutrient intakes and health status? A systematic review of empirical evidence. Obesity Reviews. 2021;22(2):e13139. doi: 10.1111/obr.13139.

34. Vandevijvere S, Vanderlee L. Effect of Formulation, Labelling, and Taxation Policies on the Nutritional Quality of the Food Supply. Current Nutrition Reports. 2019;8(3):240–9. doi: 10.1007/s13668-019-00289-x.

35. Pan American Health Organization. Plan of Action for the Elimination of Industrially Produced Trans-Fatty Acids 2020-2025. Washington, D.C.: 2020.

36. Decreto Supremo N.° 033-2016-SA – Reglamento que establece el proceso de reducción gradual hasta la eliminación de las grasas trans en los alimentos y bebidas no alcohólicas procesados industrialmente., (2016).

37. Downs SM, Thow AM, Leeder SR. The effectiveness of policies for reducing dietary trans fat: a systematic review of the evidence. Bull World Health Organ. 2013;91(4):262–9h. Epub 20130204. doi: 10.2471/blt.12.111468. PubMed PMID: 23599549; PubMed Central PMCID: PMC3629452.

38. Rebolledo N, Bercholz M, Corvalán C, Ng SW, Taillie LS. Did the sweetness of beverages change with the Chilean Food Labeling and Marketing Law? A before and after study. Frontiers in Nutrition. 2022;9. doi: 10.3389/fnut.2022.1043665.

39. Zancheta Ricardo C, Corvalán C, Smith Taillie L, Quitral V, Reyes M. Changes in the Use of Non-nutritive Sweeteners in the Chilean Food and Beverage Supply After the Implementation of the Food Labeling and Advertising Law. Frontiers in Nutrition. 2021;8. doi: 10.3389/fnut.2021.773450.

40. Rose G. Sick individuals and sick populations. Int J Epidemiol. 1985;14(1):32–8. doi: 10.1093/ije/14.1.32. PubMed PMID: 3872850.

41. Løvhaug AL, Granheim SI, Djojosoeparto SK, Harrington JM, Kamphuis CBM, Poelman MP, et al. The potential of food environment policies to reduce socioeconomic inequalities in diets and to improve healthy diets among lower socioeconomic groups: an umbrella review. BMC Public Health. 2022;22(1). doi: 10.1186/s12889-022-12827-4.

42. Scrinis G, Monteiro CA. Ultra-processed foods and the limits of product reformulation. Public Health Nutrition. 2018;21(1):247–52. doi: 10.1017/s1368980017001392.

43. Diez-Canseco F, Najarro L, Cavero V, Saavedra-Garcia L, Taillie LS, Carpentier FRD, et al. Recall, understanding, use, and impact of front-of-package warning labels on ultra-processed foods: A qualitative study with mothers of preschool children in Peru. PLOS Global Public Health. 2024;4(12):e0003938. doi: 10.1371/journal.pgph.0003938.

44. Popkin BM, Barquera S, Corvalan C, Hofman KJ, Monteiro C, Ng SW, et al. Towards unified and impactful policies to reduce ultra-processed food consumption and promote healthier eating. The Lancet Diabetes & Endocrinology. 2021;9(7):462–70. doi: 10.1016/S2213-8587(21)00078-4.

45. Meza-Hernández M, Yabiku-Soto K, Saavedra-Garcia L, Diez-Canseco F. Declaración de información nutricional en el etiquetado de bebidas y alimentos procesados y ultraprocesados ofertados en una cadena de supermercados de Lima en el 2022. Revista Peruana de Medicina Experimental y Salud Pública. 2023:141–9. doi: 10.17843/rpmesp.2023.402.12714.

46. Meza-Hernández M, Yabiku-Soto K, Saavedra-Garcia L, Diez-Canseco F. El tamaño del empaque de las galletas ultraprocesadas evita que lleven octógonos a pesar de tener alto contenido de azúcar y grasas saturadas. Revista Peruana de Medicina Experimental y Salud Pública. 2023:369–71. doi: 10.17843/rpmesp.2023.403.13119.

