## Supporting Information for "Peru’s front-of-package warning label policy and changes in the healthfulness of the food supply: A pre-post observational study"

**S1 Table.** Cutoffs of the Promotion of Healthy Eating for Boys, Girls, and Adolescents law (1, 2).

| Nutrient of Concern | Product | First Phase of the FOPWL Policy (June 2019) | Second Phase of the FOPWL Policy (September 2021) |
|---|---|---|---|
| Total sugars | Foods | Greater than or equal to 22.5 g/100 g | Greater than or equal to 10 g/100 g |
| | Beverages | Greater than or equal to 6 g/100 mL | Greater than or equal to 5 g/100 mL |
| Sodium | Foods | Greater than or equal to 800 mg/100 g | Greater than or equal to 400 mg/100 g |
| | Beverages | Greater than or equal to 100 mg/100 mL | Greater than or equal to 100 mg/100 mL |
| Saturated Fats | Foods | Greater than or equal to 6 g/100 g | Greater than or equal to 4 g/100 g |
| | Beverages | Greater than or equal to 3 g/100 mL | Greater than or equal to 3 g/100 mL |
| Trans Fats | Foods | Contains trans fats | Added trans fats prohibited |
| | Beverages | | |

FOPWL: front-of-package warning label.

**S2 Table.** Type of foods classified in each food or beverage group.

| Group | Study Categories | Descriptions/ Examples |
|---|---|---|
| **Foods** | Bread and Savory Bakery | Bread, salty crackers, other savory bakery, flour tortillas |
| | Breakfast Cereals and Bars | Breakfast cereals, cereal bars |
| | Dairy Based Products | Cheese, cream |
| | Candies, Sweet Confectionery, and Processed Fruit | Candies without chocolate, chewables, chocolate candies, gelatin desserts, other sweets, conserved canned fruit |
| | Ice creams | Milk-based ice creams, water-based ice creams |
| | Nuts and Savory Snacks | Corn based chips, nuts and seeds, popcorn, potato chips, puffed snacks, other savory snacks |
| | Ready to Eat Meals | Ready-to-eat preparations, instant soups, creamy vegetable soup |
| | Processed Meats, Fish, and Seafood | Processed meats, ham, sausages, processed fish, processed seafood |
| | Savory Sauces, Spreads, and Fats | Salad dressings, Ketchup, tomato pastes, mustard, chili sauces, BBQ sauces, mayonnaise, margarine, spreadable fats |
| | Sweet Bakery | Cookies, sweet crackers, sweet pastries, cake, waffers |
| | Sweet Spreads | Jam, caramel sauce, marmalades, peanut butter and jelly, chocolate spreads |
| | Yogurt* | Regular and light yogurt, drinkable yogurt |
| **Beverages** | Sugar-sweetened beverages | Carbonated drinks, juices and nectars, energy drinks, sports drinks, coffee and teas |
| | Milk and Milk-Based Drinks | Milk, flavored milks, powdered milks, dairy drink mixes |
| | Milk Substitutes and Formulas | Almond-based drinks, soy-based drinks, coconut-based drinks, formulas and supplements |

*Yogurts were classified under food categories because their nutritional information is reported in grams rather than milliliters.

**S3 Table.** Frequency and proportion of products by food or beverage group in T0, T1, and the longitudinal sample.

| | **Pre-implementation cross-sectional sample (T0)** | | **Post-implementation cross-sectional sample (T1)** | | **Longitudinal sub-sample** | |
|---|---|---|---|---|---|---|
| | N | % | N | % | N | % |
| **Total Foods** | 1,839 | | 2,383 | | 1,279 | |
| Bread and Other Bakery | 146 | 5.9 | 178 | 5.9 | 123 | 7.3 |
| Cereals | 159 | 6.4 | 205 | 6.8 | 117 | 6.9 |
| Dairy Based Products | 62 | 2.5 | 99 | 3.3 | 41 | 2.4 |
| Candies, Sweet Confectionery, and Processed Fruit | 493 | 19.9 | 575 | 19.1 | 331 | 19.5 |
| Ice Creams | 45 | 1.8 | 70 | 2.3 | 32 | 1.9 |
| Nuts and Savory Snacks | 170 | 6.9 | 205 | 6.8 | 108 | 6.4 |
| Ready-to-Eat Meals | 89 | 3.6 | 141 | 4.7 | 62 | 3.7 |
| Processed Meats, Fish, and Seafood | 103 | 4.2 | 186 | 6.2 | 75 | 4.4 |
| Savory Sauces, Spreads, and Fats | 179 | 7.2 | 189 | 6.3 | 108 | 6.4 |
| Sweet Bakery | 210 | 8.5 | 293 | 9.7 | 153 | 9.0 |
| Sweet Spreads | 65 | 2.6 | 89 | 3.0 | 45 | 2.7 |
| Yogurt | 118 | 4.8 | 153 | 5.1 | 84 | 5.0 |
| **Total Beverages** | 642 | | 635 | | 415 | |
| Sugar-sweetened Beverages | 442 | 17.8 | 398 | 13.2 | 269 | 15.9 |
| Milk and Milk-based Drinks | 127 | 5.1 | 148 | 4.9 | 99 | 5.8 |
| Milk Substitutes and Formulas | 73 | 2.9 | 89 | 3.0 | 47 | 2.8 |
| **Total** | 2,481 | | 3,018 | | 1,694 | |

**S4 Table**. Cross-sectional changes in the proportion of "high in" nutrients of concern (or any high in) before (T0) and after the first phase of Peru's Law (T1) for overall products.

| | 2019 ($T_0$) % (95% CI) | 2020 ($T_1$) % (95% CI) | Difference (T0 vs T1) Absolute change | Relative change (% of T0) | p-value |
|---|---|---|---|---|---|
| **Overall** | n = 2,481 | n = 3,018 | | | |
| Any "High in" | 54.5 (52.5, 56.5) | 48.7 (47.0, 50.5) | –5.7 | –10.6 | **<0.01** |
| High in Sugars | 38.5 (36.6, 40.5) | 33.3 (31.6, 35.0) | –5.3 | –13.6 | **<0.01** |
| High in Sodium | 7.9 (6.9, 9.0) | 6.9 (6.0, 7.8) | –1.0 | –13.2 | 0.14 |
| High in Saturated Fats | 28.8 (27.0, 30.6) | 29.2 (27.6, 30.9) | 0.5 | 1.6 | 0.71 |
| Contains Trans Fats | 6.4 (5.4, 7.4) | 4.1 (3.4, 4.8) | –2.3 | –35.4 | **<0.01** |

Values represent the sample size and the proportion of regulated products.
Cutoffs correspond to the limits on the amount of energy or nutrient of concern for the implementation of the first phase of the law (i.e., for solids, per 100g: 22.5 g of sugars, 6 g of saturated fats, 800 mg of sodium; for liquids, per 100 mL: 6 g of sugars, 3 g of saturated fats, 100 mg of sodium).
T0: pre-implementation period, March to May 2019 (n = 2,481); T1: post-implementation of the 1st phase of the law, March 2020 to February 2021 (n = 3,018).

**S5 Table**. Cross-sectional changes in the proportion of "high in" nutrients of concern (or any high in) before (T0) and after the first phase of Peru's Law (T1) for sugar sweetened beverages subgroups.

| | 2019 ($T_0$) % (95% CI) | 2020 ($T_1$) % (95% CI) | Difference (T0 vs T1) Absolute change | Relative change (% of T0) | p-value |
|---|---|---|---|---|---|
| **Carbonated drinks** | N= 102 | N= 109 | | | |
| Any "High in" | 49.0 (39.2, 58.7) | 39.4 (30.3, 48.6) | –9.6 | –19.5 | 0.16 |
| High in Sugars | 49.0 (39.3, 58.7) | 39.4 (30.3, 48.6) | –9.6 | –19.5 | 0.16 |
| High in Sodium | 0 | 0 | NA | NA | NA |
| **Juices and Nectars** | N = 165 | N = 196 | | | |
| Any "High in" | 42.4 (34.9, 50.0) | 13.3 (8.5, 18.0) | –29.2 | –68.7 | **<0.01** |
| High in Sugars | 41.6 (34.0, 49.2) | 12.2 (7.7, 16.8) | –29.4 | –70.6 | **<0.01** |
| High in Sodium | 0.2 (0.0, 3.9) | 0.1 (0.0, 2.4) | –0.1 | –43.9 | 0.53 |
| **Sports and energy drinks** | N = 55 | N = 39 | | | |
| Any "High in" | 30.9 (18.7, 43.1) | 15.4 (4.1, 26.7) | –15.5 | –50.2 | 0.07 |
| High in Sugars | 30.9 (18.7, 43.1) | 15.4 (4.1, 26.7) | –15.5 | –50.2 | 0.07 |
| High in Sodium | 0 | 0 | NA | NA | NA |
| **Coffee and teas** | N = 120 | N = 55 | | | |
| Any "High in" | 5.8 (1.6, 10.0) | 3.6 (0.0, 8.6) | –2.2 | –37.7 | 0.51 |
| High in Sugars | 5.8 (1.6, 10.0) | 3.6 (0.0, 8.6) | –2.2 | –37.7 | 0.51 |
| High in Sodium | 0 | 0 | NA | NA | NA |

Values represent the sample size and the proportion of regulated products.
Cutoffs correspond to the limits on the amount of energy or nutrient of concern for the implementation of the first phase of the law (i.e., per 100 mL: 6 g of sugars, 3 g of saturated fats, 100 mg of sodium).
T0: pre-implementation period, March to May 2019; T1: post-implementation of the 1st phase of the law, March 2020 to February 2021.

**S6 Table**. Longitudinal changes in the proportion of "high in" nutrients of concern (or any high in) before (T0) and after the first phase of Peru's Law (T1) for overall products.

| | **2019 ($T_0$) % (95% CI)** | **2020 ($T_1$) % (95% CI)** | **Difference (T0 vs T1)** | | |
|---|---|---|---|---|---|
| | | | **Absolute change** | **Relative change (% of T0)** | **p-value** |
| **Overall** | N = 1,694 | | | | |
| Any "High in" | 55.1 (52.7, 57.4) | 47.6 (45.3, 50.0) | –7.4 | –13.5 | **<0.01** |
| High in Sugars | 39.9 (37.6, 42.3) | 33.9 (31.7, 36.2) | –6.0 | –15.0 | **<0.01** |
| High in Sodium | 7.7 (6.4, 9.0) | 6.7 (5.5, 7.9) | –1.0 | -13.0 | **<0.01** |
| High in Saturated Fats | 28.1 (26.0, 30.3) | 26.0 (23.9, 28.1) | –2.1 | –7.3 | **<0.01** |
| Contains Trans Fats | 6.3 (5.2, 7.5) | 4.8 (3.8, 5.9) | –1.5 | –24.1 | **<0.01** |

Values represent the sample size and the proportion of regulated products.
Cutoffs correspond to the limits on the amount of energy or nutrient of concern for the implementation of the first phase of the law (i.e., for solids, per 100g: 22.5 g of sugars, 6 g of saturated fats, 800 mg of sodium; for liquids, per 100 mL: 6 g of sugars, 3 g of saturated fats, 100 mg of sodium).
T0: pre-implementation period, March to May 2019 (n = 1,694); T1: post-implementation of the 1st phase of the law, March 2020 to February 2021 (n = 1,694).

**S7 Table**. Changes in the proportion of "high in" nutrients of concern (or any high in) before (T0) and after the first phase of Peru's Law (T1) by food and beverage group, longitudinal analysis.

| | 2019 ($T_0$) % (95% CI) | 2020 ($T_1$) % (95% CI) | Difference (T0 vs T1) Absolute | Relative (% of T0) | p-value |
|---|---|---|---|---|---|
| **Food Groups** | | | | | |
| **Bread and Savory Bakery** | N = 123 | | | | |
| Any "High in" | 18.7 (11.8, 25.6) | 13.0 (7.0, 19.0) | –5.7 | –30.4 | **0.01** |
| High in Sugars | 0 | 0 | NA | NA | NA |
| High in Sodium | 10.6 (5.1, 16.0) | 6.5 (2.1, 10.9) | –4.1 | –38.5 | 0.06 |
| High in Saturated Fats | 11.4 (5.7, 17.0) | 5.7 (1.6, 9.8) | –5.7 | –50.0 | **0.01** |
| Contains Trans Fats | 2.4 (–0.3, 5.2) | 1.6 (–0.6, 3.9) | –0.8 | –33.3 | 0.32 |
| **Breakfast Cereals and Bars** | N = 117 | | | | |
| Any "High in" | 65.0 (56.3, 73.6) | 43.6 (34.6, 52.6) | –21.4 | –32.9 | **<0.01** |
| High in Sugars | 63.8 (55.0, 72.3) | 43.1 (34.1, 52.2) | –20.7 | –32.4 | **<0.01** |
| High in Sodium | 0 | 0 | NA | NA | NA |
| High in Saturated Fats | 6.0 (1.7, 10.3) | 1.7 (0.0, 4.1) | –4.3 | –71.4 | **0.02** |
| Contains Trans Fats | 10.3 (7.7, 15.8) | 11.1 (5.4, 16.8) | 0.9 | 8.3 | 0.82 |
| **Dairy Based products*** | N = 41 | | | | |
| Any "High in" | 87.8 (77.7, 97.9) | 97.6 (92.8, 102.3) | 9.6 | 11.1 | **0.04** |
| High in Sugars | 2.4 (–2.3, 7.2) | 2.6 (–2.5, 7.6) | 0.1 | 5.1 | 0.41 |
| High in Sodium | 22.0 (9.1, 34.8) | 14.6 (3.7, 25.6) | –7.3 | –33.3 | 0.26 |
| High in Saturated Fats | 82.9 (71.3, 94.6) | 97.6 (92.8, 102.3) | 14.6 | 17.6 | **0.01** |
| Contains Trans Fats | Excluded | Excluded | NA | NA | NA |
| **Candies, Sweet Confectionery, and Processed Fruit** | N = 331 | | | | |
| Any "High in" | 78.2 (73.8, 82.7) | 78.5 (74.1, 83.0) | 0.3 | 0.4 | 0.32 |
| High in Sugars | 75.5 (70.9, 80.2) | 75.2 (70.5, 79.8) | –0.4 | –0.5 | 0.44 |
| High in Sodium | 2.1 (0.6, 3.7) | 2.1 (0.6, 3.6) | 0 | 0 | 1.00 |
| High in Saturated Fats | 52.0 (46.6, 57.4) | 52.6 (47.2, 58.0) | 0.6 | 1.2 | 0.10 |
| Contains Trans Fats | 9.7 (6.5, 12.9) | 8.8 (5.7, 11.8) | –0.9 | –9.4 | 0.44 |
| **Ice creams** | N = 32 | | | | |
| Any "High in" | 78.1 (63.6, 92.7) | 62.5 (45.5, 79.5) | –15.6 | –20.0 | **0.02** |
| High in Sugars | 56.3 (38.8, 73.7) | 37.5 (20.5, 54.5) | –18.8 | –33.3 | **0.01** |
| High in Sodium | 0 | 0 | NA | NA | NA |
| High in Saturated Fats | 56.3 (38.8, 73.7) | 56.3 (38.8, 73.7) | 0 | 0 | 1.0 |
| Contains Trans Fats | 0 | 0 | NA | NA | NA |
| **Nuts and savory snacks** | N = 108 | | | | |
| Any "High in" | 63.9 (54.8, 73.0) | 59.3 (49.9, 68.6) | –4.6 | –7.2 | 0.06 |
| High in Sugars | 7.5 (2.5, 12.5) | 6.5 (1.8, 11.2) | –0.9 | –12.5 | 0.32 |
| High in Sodium | 19.4 (11.9, 26.9) | 19.4 (11.9, 26.9) | 0 | 0 | 1.00 |
| High in Saturated Fats | 48.1 (38.7, 57.6) | 46.3 (36.8, 55.7) | –1.9 | –3.8 | 0.32 |
| Contains Trans Fats | 0.9 (–0.9, 2.7) | 2.8 (–0.3, 5.9) | 1.9 | 200.0 | 0.16 |
| **Ready-to-eat meals** | N = 62 | | | | |
| Any "High in" | 29.0 (17.6, 40.4) | 16.1 (6.9, 25.4) | –12.9 | –44.4 | **0.02** |
| High in Sugars | 0 | 0 | NA | NA | NA |
| High in Sodium | 0 (0, 5.8) | 6.5 (1.8, 15.7) | 6.5 | NA | 0.07 |
| High in Saturated Fats | 17.7 (8.2, 27.3) | 6.5 (0.3, 12.6) | –11.3 | –63.6 | **0.01** |
| Contains Trans Fats | 19.4 (9.4, 29.3) | 9.7 (2.3, 17.1) | –9.7 | –50.0 | 0.05 |

| | | | | | |
|---|---|---|---|---|---|
| **Processed meats, fish, and seafood** | N = 75 | | | | |
| Any "High in" | 40.0 (28.8, 51.2) | 34.7 (23.8, 45.5) | –5.3 | –13.3 | 0.10 |
| High in Sugars | 0 | 0 | NA | NA | NA |
| High in Sodium | 38.7 (27.6, 49.8) | 33.3 (22.6, 44.1) | –5.3 | –13.8 | 0.10 |
| High in Saturated Fats | 15.7 (6.6, 22.7) | 17.3 (8.7, 26.6) | 2.7 | 18.2 | 0.15 |
| Contains Trans Fats | 4.0 (–0.5, 8.5) | 2.7 (–1.0, 6.3) | –1.3 | –33.3 | 0.57 |
| **Savory sauces, spreads, and fats** | N = 108 | | | | |
| Any "High in" | 65.7 (56.7, 74.7) | 57.4 (48.0, 66.8) | –8.3 | –12.7 | **<0.01** |
| High in Sugars | 12.6 (6.2, 19.1) | 12.6 (6.2, 19.1) | 0 | 0 | 1.0 |
| High in Sodium | 41.7 (32.3, 51.0) | 34.6 (25.5, 43.6) | –7.1 | –17.0 | **<0.01** |
| High in Saturated Fats | 31.1 (22.3, 40.0) | 28.7 (20.1, 37.3) | –2.4 | –7.8 | 0.08 |
| Contains Trans Fats | 8.3 (3.1, 13.6) | 8.3 (3.1, 13.6) | 0 | 0 | 1.0 |
| **Sweet bakery** | N = 153 | | | | |
| Any "High in" | 90.2 (85.5, 94.9) | 79.7 (73.3, 86.1) | –10.5 | –11.6 | **<0.01** |
| High in Sugars | 81.3 (75.1, 87.6) | 71.7 (64.5, 78.9) | –9.6 | –11.8 | **<0.01** |
| High in Sodium | 0 | 0 | NA | NA | NA |
| High in Saturated Fats | 73.5 (66.4, 80.6) | 59.2 (51.4, 67.0) | –14.3 | –19.5 | **<0.01** |
| Contains Trans Fats | 22.2 (15.6, 28.8) | 9.8 (5.1, 14.5) | –12.4 | –55.9 | **<0.01** |
| **Sweet spreads** | N = 45 | | | | |
| Any "High in" | 75.6 (62.9, 88.3) | 62.2 (47.9, 76.5) | –13.3 | –17.6 | 0.05 |
| High in Sugars | 75.6 (62.3, 88.9) | 59.5 (44.5, 74.5) | –16.1 | –21.3 | **0.02** |
| High in Sodium | 0 | 0 | NA | NA | NA |
| High in Saturated Fats | 17.9 (5.8, 30.1) | 18.4 (5.9, 30.9) | 0.5 | 2.6 | 0.66 |
| Contains Trans Fats | 2.2 (–2.1, 6.6) | 2.2 (–2.1, 6.6) | 0 | 0 | 1.0 |
| **Yogurt*** | N = 84 | | | | |
| Any "High in" | 1.2 (–1.1, 3.5) | 2.4 (–0.9, 5.7) | 1.2 | 100.0 | 0.32 |
| High in Sugars | 1.2 (–1.1, 3.5) | 1.2 (–1.1, 3.5) | 0 | 0 | 1.0 |
| High in Sodium | 0 | 0 | NA | NA | NA |
| High in Saturated Fats | 0 | 0 | NA | NA | NA |
| Contains Trans Fats | Excluded | Excluded | NA | NA | NA |
| **Beverages Groups** | | | | | |
| **Sugar-sweetened Beverages** | N = 269 | | | | |
| Any "High in" | 37.9 (32.1, 43.7) | 21.9 (17.0, 26.9) | –16.0 | –42.2 | **<0.01** |
| High in Sugars | 37.2 (31.4, 43.0) | 20.8 (16.0, 25.7) | –16.4 | –44.0 | **<0.01** |
| High in Sodium | 0.8 (0.0, 1.8) | 0.8 (0.0, 1.8) | 0 | 0.4 | 0.41 |
| High in Saturated Fats | 0 | 0 | NA | NA | NA |
| Contains Trans Fats | 0 | 0 | NA | NA | NA |
| **Milk and milk-based drinks** | N = 99 | | | | |
| Any "High in" | 39.4 (29.7, 49.1) | 30.3 (21.2, 39.4) | –9.1 | –23.1 | **<0.01** |
| High in Sugars | 38.4 (28.8, 48.0) | 29.3 (20.3, 38.3) | –9.1 | –23.7 | **<0.01** |
| High in Sodium | 0 | 0 | NA | NA | NA |
| High in Saturated Fats | 3.1 (–0.4, 6.5) | 3.1 (–0.4, 6.5) | 0 | 0 | 1.0 |
| Contains Trans Fats | 0 | 0 | NA | NA | NA |
| **Milk substitutes and formulas** | N = 47 | | | | |
| Any "High in" | 25.5 (12.9, 38.1) | 31.9 (18.4, 45.4) | 6.4 | 25.0 | 0.08 |
| High in Sugars | 26.7 (13.6, 39.7) | 33.3 (19.4, 47.3) | 6.7 | 25.0 | 0.08 |
| High in Sodium | 0 | 0 | NA | NA | NA |
| High in Saturated Fats | 0 | 0 | NA | NA | NA |
| Contains Trans Fats | 0 | 0 | NA | NA | NA |

Values represent the sample size and the proportion of regulated products.
Cutoffs correspond to the limits on the amount of energy or nutrient of concern for the implementation of the first phase of the law (i.e., for solids, per 100g: 22.5 g of sugars, 6 g of saturated fats, 800 mg of sodium; for liquids, per 100 mL: 6 g of sugars, 3 g of saturated fats, 100 mg of sodium).
T0: pre-implementation period, March to May 2019 (n = 1,694); T1: post-implementation of the 1st phase of the law, March 2020 to February 2021 (n = 1,702).
P-value < 0.05. Comparison between T0 and T1 were done using Estimated marginal means (EMMs) from logistic regression.
* Trans fat analysis for dairy and yogurt products was excluded, in line with Peruvian regulations that exempt the declaration of naturally occurring trans fats.

**S8 Table**. Longitudinal changes in the proportion of "high in" nutrients of concern (or any high in) before (T0) and after the first phase of Peru's Law (T1) for sugar sweetened beverages subgroups.

| | **2019 ($T_0$) % (95% CI)** | **2020 ($T_1$) % (95% CI)** | **Difference (T0 vs T1)** | | |
|---|---|---|---|---|---|
| | | | **Absolute change** | **Relative change (% of T0)** | **p-value** |
| **Carbonated drinks** | N= 83 | | | | |
| Any "High in" | 45.8 (35.0, 56.6) | 43.4 (32.6, 54.1) | –2.4 | –5.3 | 0.16 |
| High in Sugars | 45.8 (35.0, 56.6) | 43.4 (32.6, 54.1) | –2.4 | –5.3 | 0.16 |
| High in Sodium | 0 | 0 | NA | NA | NA |
| **Juices and Nectars** | N = 113/114 | | | | |
| Any "High in" | 43.4 (34.2, 52.5) | 14.0 (7.6, 20.4) | –29.3 | –67.6 | **<0.01** |
| High in Sugars | 41.6 (32.5, 50.7) | 12.3 (6.2, 18.3) | –29.3 | –70.5 | **<0.01** |
| High in Sodium | 1.8 (0.0, 4.2) | 1.8 (0.0, 4.2) | 0 | 0 | 1.0 |
| **Sports and energy drinks** | N = 34 | | | | |
| Any "High in" | 29.4 (13.9, 45.0) | 14.7 (2.6, 26.8) | –14.7 | –50.0 | **0.02** |
| High in Sugars | 29.4 (13.9, 45.0) | 14.7 (2.6, 26.8) | –14.7 | –50.0 | **0.02** |
| High in Sodium | 0 | 0 | NA | NA | NA |
| **Coffee and teas** | N = 39 | | | | |
| Any "High in" | 12.8 (2.2, 23.5) | 5.1 (0.0, 12.1) | –7.7 | –60.0 | 0.18 |
| High in Sugars | 12.8 (2.2, 23.5) | 2.6 (0.0, 7.6) | –10.3 | –80.0 | **0.04** |
| Contains trans fat | 0 | 0.0 | –0.0 | –0.0 | 0.32 |

Values represent the sample size and the proportion of regulated products.
Cutoffs correspond to the limits on the amount of energy or nutrient of concern for the implementation of the first phase of the law (i.e., per 100 mL: 6 g of sugars, 3 g of saturated fats, 100 mg of sodium).
T0: pre-implementation period, March to May 2019; T1: post-implementation of the 1st phase of the law, March 2020 to February 2021.

**S9 Table**. Changes in quartiles of nutrients of concern before (T0) and after first phase of Peru's Law (T1) for overall food and beverages and by food and beverage group, longitudinal analysis.

| | **TOTAL SUGARS (g/100g-mL)** | | **SODIUM (g/100g-mL)** | | **SATURATED FATS (mg/100g-mL)** | |
|---|---|---|---|---|---|---|
| | **2019 ($T_0$)** | **2020 ($T_1$)** | **2019 ($T_0$)** | **2020 ($T_1$)** | **2019 ($T_0$)** | **2020 ($T_1$)** |
| Overall Foods | | | | | | |
| p25 | 3.0 | 3.0 | 60.0 | 60.0 | 0.1 | **0.4** |
| p50 | 12.0 | 12.0 | 255.8 | 245.6 | 3.0 | 3.1 |
| p75 | 37.5 | 37.2 | 472.0 | 460.0 | 10.5 | 10.0 |
| Overall Beverages | | | | | | |
| p25 | 3.6 | 3.7 | 7.5 | 7.9 | 0 | 0 |
| p50 | 5.7 | 5.3 | 23.0 | 25.0 | 0 | 0 |
| p75 | 8.3 | 5.9 | 46.5 | 49.9 | 0.9 | 0.9 |
| Food Groups | | | | | | |
| Bread and Savory Bakery | | | | | | |
| T0 cutoff: | NA | | 88th percentile | | 89th percentile | |
| p25 | 2.8 | 2.8 | 339.0 | 364.8 | 0.6 | **0.9** |
| p50 | 4.0 | 4.4 | 426.7 | 440 | 2.0 | 2.0 |
| p75 | 6.3 | 7.0 | 700.0 | 700.0 | 3.8 | 3.8 |
| Breakfast Cereals and Bars | | | | | | |
| T0 cutoff: | 37th percentile | | 98th percentile | | 97th percentile | |
| p25 | 17.0 | 15.0 | 160.0 | 148.6 | 0 | 0 |
| p50 | 26.3 | **21.8** | 302.0 | 309.5 | 0.5 | 0.7 |
| p75 | 32.2 | 30.2 | 426.0 | 416.7 | 2.4 | 2.3 |
| Dairy Based Products | | | | | | |
| T0 cutoff: | 99th percentile | | 78th percenile | | 10th percentile | |
| p25 | 0.7 | 0 | 50.0 | 53.7 | 11.0 | 11.7 |
| p50 | 2.7 | 2.5 | 550.0 | **440.0** | 15.9 | 16.5 |
| p75 | 3.3 | 4.0 | 781.0 | 720.0 | 19.3 | 20.0 |
| Candies, Sweet Confectionery, and Processed Fruit | | | | | | |
| T0 cutoff: | 24th percentile | | 98th percentile | | 47th percentile | |
| p25 | 26.7 | **20.0** | 19.6 | 22.2 | 0 | 0 |
| p50 | 49.6 | **47.6** | 51.8 | 52.0 | 8.5 | 8.8 |
| p75 | 57.0 | 56.9 | 92.6 | 94.7 | 18.0 | 18.0 |
| Ice Cream | | | | | | |
| T0 cutoff: | 44th percentile | | NA | | 44th percentile | |
| p25 | 21.2 | 21.2 | 63.0 | 58.4 | 3.6 | 3.6 |
| p50 | 23.3 | 21.8 | 72.0 | 72.0 | 8.2 | 8.2 |
| p75 | 24.0 | 24.1 | 85.0 | 81.0 | 10.3 | 10.3 |
| Nuts and Savory Snacks | | | | | | |
| T0 cutoff: | 91st percentile | | 81st percentile | | 53rd percentile | |
| p25 | 0.4 | 0.7 | 339.3 | 320.0 | 2.9 | 2.8 |
| p50 | 3.0 | 3.0 | 460.0 | 440.0 | 6.0 | 5.4 |
| p75 | 6.0 | 6.7 | 710.0 | 710.0 | 10.0 | 10.0 |
| Ready-To-Eat Meals | | | | | | |
| T0 cutoff: | NA | | NA | | 92nd percentile | |
| p25 | 0 | 0.1 | 300.0 | 293.2 | 0.6 | 0.6 |
| p50 | 0.6 | 0.8 | 385.2 | 373.5 | 1.4 | 1.3 |
| p75 | 1.5 | 2.4 | 494.1 | 513.9 | 3.0 | 3.8 |
| Processed Meats, Fish, and Seafood | | | | | | |
| T0 cutoff: | NA | | 61st percentile | | 84th percentile | |
| p25 | 0 | 0 | 384.0 | 361.5 | 0.6 | 0.8 |
| p50 | 0 | 0 | 602.7 | 600.0 | 1.8 | 2.7 |
| p75 | 0.7 | 0.8 | 1100.0 | **1000.0** | 4.3 | 4.3 |

| | | | | | | |
|---|---|---|---|---|---|---|
| Savory Sauces, Spreads, and Fats | | | | | | |
| T0 cutoff: | 89th percentile | | 59st percentile | | 68th percentile | |
| p25 | 0 | 0 | 520.0 | **445.0** | 0 | 0 |
| p50 | 3.3 | 3.1 | 700.0 | 678.6 | 0.5 | 1.0 |
| p75 | 8.0 | 7.1 | 1062.5 | **964.3** | 7.0 | 7.0 |
| Sweet Bakery | | | | | | |
| T0 cutoff: | 19th percentile | | NA | | 28th percentile | |
| p25 | 23.5 | 22.3 | 160.0 | 160.0 | 5.8 | **5.5** |
| p50 | 30.2 | 30.2 | 255.8 | 254.5 | 9.2 | 8.3 |
| p75 | 38.1 | 38.1 | 361.7 | 355.6 | 12.0 | 11.9 |
| Sweet Spreads | | | | | | |
| T0 cutoff: | 23rd percentile | | NA | | 82nd percentile | |
| p25 | 23.5 | 18.8 | 15.0 | 15.8 | 0 | 0 |
| p50 | 40.0 | 40.0 | 44.0 | 40.0 | 0 | 0 |
| p75 | 55.5 | 54.1 | 125.0 | 115.0 | 5.2 | 5.2 |
| Yogurt | | | | | | |
| T0 cutoff: | NA | | NA | | NA | |
| p25 | 5.7 | 8.1 | 50.0 | 50.0 | 0 | 0 |
| p50 | 10.0 | 10.0 | 55.2 | 55.8 | 1.0 | 1.0 |
| p75 | 12.0 | 12.0 | 74.0 | 74.0 | 1.4 | 1.4 |
| Beverages Groups | | | | | | |
| Sugar-sweetened beverages | | | | | | |
| T0 cutoff: | 65th percentile | | 99th percentile | | NA | |
| p25 | 0.4 | 0.4 | 5.8 | 6.0 | 0 | 0 |
| p50 | 5.8 | 5.4 | 9.0 | 10.0 | 0 | 0 |
| p75 | 8.0 | **5.9** | 22.6 | 23.3 | 0 | 0 |
| Milk and Milk-Based Drinks | | | | | | |
| T0 cutoff: | 58th percentile | | 96th percentile | | 97th percentile | |
| p25 | 4.8 | 4.8 | 45.6 | 49.9 | 1.0 | 1.0 |
| p50 | 5.8 | 5.0 | 51.0 | 56.1 | 1.4 | 1.3 |
| p75 | 9.0 | 8.4 | 57.0 | 67.0 | 2.0 | 2.0 |
| Milk Substitutes and Formulas | | | | | | |
| T0 cutoff: | 64th percentile | | 96th percentile | | NA | |
| p25 | 3.0 | 3.0 | 36.0 | 38.0 | 0.1 | 0.2 |
| p50 | 5.0 | 5.0 | 39.9 | 47.0 | 0.5 | 0.7 |
| p75 | 7.0 | 7.0 | 50.0 | 56.0 | 1.0 | 1.1 |

T0: pre-implementation period, March to May 2019 (n = 1,702); T1: post-implementation of the 1st phase of the law, March 2020 to February 2021 (n = 1,694).
Quartiles and p-values were obtained from quantile regression models (one model per nutrient per food or beverage group), using the implementation period as independent variable. Significant p-values are bold and represent a p-value <0.05 versus T0.

### References

1. Manual de Advertencias Publicitarias en el marco de lo establecido en la Ley Nº 30021, Ley de promoción de la alimentación saludable para niños, niñas y adolescentes, y su Reglamento aprobado por Decreto Supremo Nº 017-2017-SA, Stat. DECRETO SUPREMO Nº 012-2018-SA (2018).
2. Ley de Promoción de la Alimentación Saludable para Niños, Niñas y Adolescentes, Stat. LEY No 30021 (17 de Mayo, 2013, 2013).
